# Empirical Calibration of a Simulation Model of Opioid Use Disorder

**DOI:** 10.1101/2022.01.24.22269191

**Authors:** R. W. M. A. Madushani, Jianing Wang, Michelle Weitz, Benjamin Linas, Laura F. White, Stavroula Chrysanthopoulou

**Affiliations:** Boston Medical Center and Boston University School of Medicine, Section of Infectious Diseases, Department of Medicine, 801 Massachusetts Ave, 2nd Floor, Boston, MA 02118; Department of Biostatistics, Boston University School of Public Health, 715 Albany Street, Boston, MA 02118; Department of Biostatistics, Brown University School of Public Health, 121 S Main St, Providence, RI 02912

**Author notes:** **Corresponding Author:** R. W. M. A. Madushani, PhD, 801 Massachusetts Ave, Crosstown Center, Boston, MA, 02118.

## Abstract

Simulation models of opioid use disorder (OUD) aim at evaluating the impact of different treatment strategies on population-level outcomes. Researching Effective Strategies to Prevent Opioid Death (RESPOND) is a dynamic population state-transition model that simulates Massachusetts (MA) OUD population synthesizing data from the MA Public Health Data Warehouse, published survey studies, and randomized trials. We implement an empirical calibration approach to fit RESPOND to multiple calibration targets, including yearly counts of fatal overdoses and detox admissions in 2013-2015, and 2015 OUD population counts in MA. We used capture-recapture analysis to estimate the OUD population and to quantify uncertainty around calibration targets.^1^ The empirical calibration approach involves Latin hypercube sampling for a parameter search of the multidimensional space, comprising demographics of “arrivals”, overdose rates, treatment transition rates, and substance use transition probabilities. The algorithm accepts proposed parameter values when the respective model outputs are “close” to the observed calibration targets based on uncertainty ranges of targets. Calibration provided an excellent fit to the model calibration targets. The flexibility of the algorithm also allowed us to identify certain “questionable” parts of the model structure and explore the underlying relationships between the model parameters in an efficient manner. The calibrated model also provided a good fit to validation targets: non-overdose related deaths, percentage of active OUDs, and all types of overdose counts (fatal and non-fatal). In addition, the resulting set of values for the calibrated parameters will inform the priors of a more comprehensive Bayesian calibration. The calibrated RESPOND model will be employed to improve shared decision-making for OUD.

## 1. Introduction

Opioid use disorder (OUD) is an epidemic in the U.S and opioid overdose is a growing public health emergency.^2-5^ In the context of this public health crisis, policymakers and public health officials seek effective strategies to address opioid use disorder and to reduce opioid death. Buprenorphine-naloxone, naltrexone, and methadone are all FDA-approved medications for OUD (MOUDs) that decrease opioid use, mortality, and criminal activity.^6^ These treatments are under-utilized, however, fewer than 25% of people with OUD being on treatment.^7^ One of the key pillars of the U.S. response to the overdose crisis is increasing access to MOUD. Doing so, however, requires identifying and working in new venues and among new populations who are not currently engaged with treatment. The relative value of different venues, the budget impact of expanding MOUD, and the cost-effectiveness of various models of care are all essential knowledge for decision making, but they are not known.

Simulation models provide a tool for evaluating the impact of different care delivery and treatment strategies on population-level outcomes. Simulation models integrate data from multiple sources to translate outcomes from clinical studies to policy-relevant data about population health and cost. A model that simulates a state-level population of people with OUD could be a useful tool to investigate delivery system innovations and project the impact on public health outcomes and cost and will be invaluable in the fight against OUD. However, due to the complicated nature of how the system operates, such a model consists of a complex structure to capture the real-world dynamics.

In previous studies, simulation models have been used to investigate the health and economic effects of prevention, treatment, or harm reduction interventions targeting opioid misuse and/or overdose.^8^ For example, a system dynamic simulation model has been used to project effects of interventions to lower prescription opioid misuse on opioid overdose deaths in U.S.^9^, and agent-based simulation has been productively employed to estimate the impact of evidence-based strategies for preventing opioid overdose deaths in a selected number of states.^10^

In our work, with the goal of informing state-level innovation for low-barrier access to MOUDs, we developed a dynamic population, state-transition model to simulate the OUD population in Massachusetts (MA). We named the model the Researching Effective Strategies to Prevent Opioid Death (RESPOND) Model. RESPOND is built with a complex model structure that allows flexibility to study various interventions incorporating different treatments to prevent opioid overdoses. One essential challenge to modeling OUD and the overdose crisis, however, is the need to estimate parameter values for behaviors and events that are impossible to observe and/or are difficult to measure with bias-related to stigma and under-reporting of risk. The structural complexity of RESPOND combined with the scarcity of available data for opioid modeling poses a challenge to calibrating and validating the model.^11^

In this work, we present an empirical approach that was used to calibrate the RESPOND model and that could be useful to other researchers seeking to develop models focused on evaluating interventions for opioid use disorder.

## 2. Methods

### 2.1 The RESPOND model

We developed and calibrated the Researching Effective Strategies to Prevent Opioid Death (RESPOND) model, a dynamic, cohort-based, state-transition model that simulates the OUD population in Massachusetts. Figure 1 provides an overview of the RESPOND model structure. The model simulates the clinical progression of a cohort with opioid use disorder assuming five major health states: three related to treatment status, overdose, and death. Within each treatment state, there is a core simulation described with four distinct OUD states determined by injection status (active, non-active, and injection, non-injection). Fig 1 presents the set of possible transitions that RESPOND allows between the health and OUD states.

**Figure 1.**
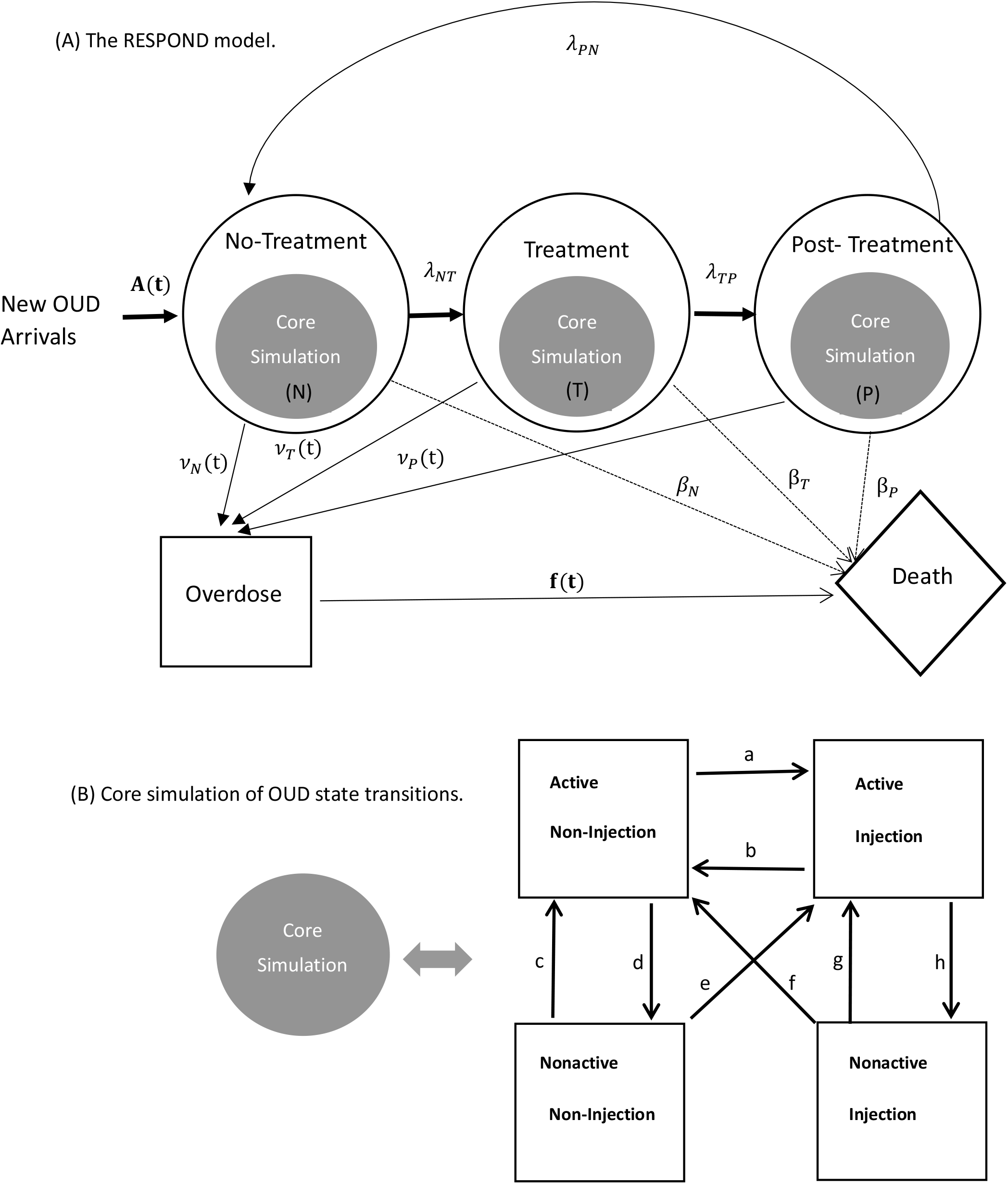
Model Structure of the RESPOND Model. (A) Overall model structure consists of three main health states: No-treatment, Treatment and Post-treatment denoted by N, T and P respectively. Both T and P are stratified by four treatment types: buprenorphine, naltrexone, methadone, and detox. **A(t)** denotes yearly time varying total new arrivals, and *λ* denotes the transition rates between health states N, T and P. *v(t)* denotes yearly time varying overdose rates parameter in health states, and **f**(**t**) denotes yearly time varying fatal overdose proportion applied on total overdoses (both fatal and non-fatal) resulted from all health states. β denotes non-overdose related other cause mortality rates in health states N, T and P represented by dashed arrows. (B) Core simulation of substance use state transitions within each health state. Eight different transition probabilities in the core simulation are denoted by a, b, c, d, e, f, g and h.

#### Core simulation of OUD

The core simulation of opioid use disorder includes 4 mutually exclusive states by the status and type of opioid use, i.e.: 1) active, non-injection (ANI), 2) non-active, after non-injection (NANI), 3) active, injection (AI), and 4) non-active, after injection (NAI). Active use states are characterized by risk of overdose and elevated healthcare utilization, with active injection use having a higher risk than active oral use. Non-active use states carry no risk of overdose. Throughout the simulation, there is bidirectional movement between active and non-active use states, simulating the relapsing and remitting nature of opioid use.

#### Simulation of treatment status

The core simulation of opioid use is embedded in the simulations of treatment status. RESPOND models three medication-based treatments for OUD: (methadone, buprenorphine, and naltrexone) and acute detoxification services (detox). The population that is engaged with a medication-based treatment experiences a net movement out of active drug use and into nonactive use states (treatment initiation effect), but there is bidirectional movement between use states among those engaged with MOUD treatment. MOUD provide an independent effect on overdose risk beyond their benefits of reducing active use.

The model simulates loss to follow-up from MOUD treatment by moving the population that disengages from MOUD treatment to the “post-treatment” health state, during which there is a high risk of relapse to active use states and consequently high risk of overdose.

#### Mortality

There are two components to mortality in RESPOND:

1. Overdose mortality – overdose rates are a function of drug use status and treatment and are stratified by age and sex. Among those who overdose, there is a conditional risk of death from that overdose.
2. Other cause mortality (Mortality attributable to drug use other than overdose death) – includes deaths from conditions such as infectious endocarditis and sepsis, as well as medical comorbidities that accrue over a lifetime. The model employs standardized mortality ratios (SMR) that are stratified by sex and drug use status reflecting elevated mortality among drug users to age-sex stratified actuarial lifetables of the U.S.

#### Cohort Initiation

The RESPOND model has a weekly cycle. Every week, a new population “arrives” to the simulation (no-treatment state), with independent rates of entry stratified by age and sex. The rates of arrival represent the development of new OUD cases and emigration to the state of Massachusetts. The only exit from the simulation is mortality. The balance of arrivals and deaths results in the total population size of OUD in Massachusetts.

The RESPOND model has been used in a simulation study aimed at evaluating MOUD approaches for decreasing the risk of opioid overdose.^12^

### 2.2 Data

The primary data source for RESPOND is the Massachusetts Public Health Data Repository (MA PHD). MA PHD is a longitudinally linked, administrative records database that includes service encounter data from over 16 agencies in the Commonwealth of Massachusetts. The database includes approximately 97% of the Massachusetts population, and data across agencies is linkable at the person-level. This database informed the calibration targets (Table 1) and several model input parameters (Table 2). We also use data from National Survey on Drug Use and Health (NSDUH), and US Census 2010 to inform population counts of the initial cohort and demographic proportions of new OUD arrivals. We used published studies of the natural history of OUD to estimate transitions between OUD states without treatment. National Institute on Drug Abuse (NIDA) Clinical Trials Network (CTN) studies that collected weekly urine toxicology data informs substance use transitions while in treatment. We use data from National Vital Statistics System and 2010 US Census, combined with the MA PHD, to estimate non-overdose related death rates. The RESPOND model has been deemed non-human subjects research by the Boston University/Boston Medical Center Institutional Review Board (H-38867).

**Table 1.** Calibration Targets.

| **Year** | **Total OUD Count<sup>\*,&</sup> (95% CI)** | **Number of Fatal Overdoses<sup>\*\*</sup>** | **Number of Admissions to Detox<sup>\*\*</sup>** |
| --- | --- | --- | --- |
| 2013 | ---<sup>***</sup> | 900 | 39635 |
| 2014 | ---<sup>***</sup> | 1294 | 41229 |
| 2015 | 275070 (272707, 277402) | 1562 | 38329 |

\* Total OUD Count = Alive OUD + Fatal Overdoses + Other Deaths
\*\* These are observed counts from MA PHD with no known uncertainty around the values. Therefore, we use within 10% of the values as the uncertainty interval for the empirical calibration
\*\*\* Estimates for these years were not used due to missing an important data piece in MA PHD
& Estimated from capture-recapture analysis.<sup>1</sup>

**Table 2.**
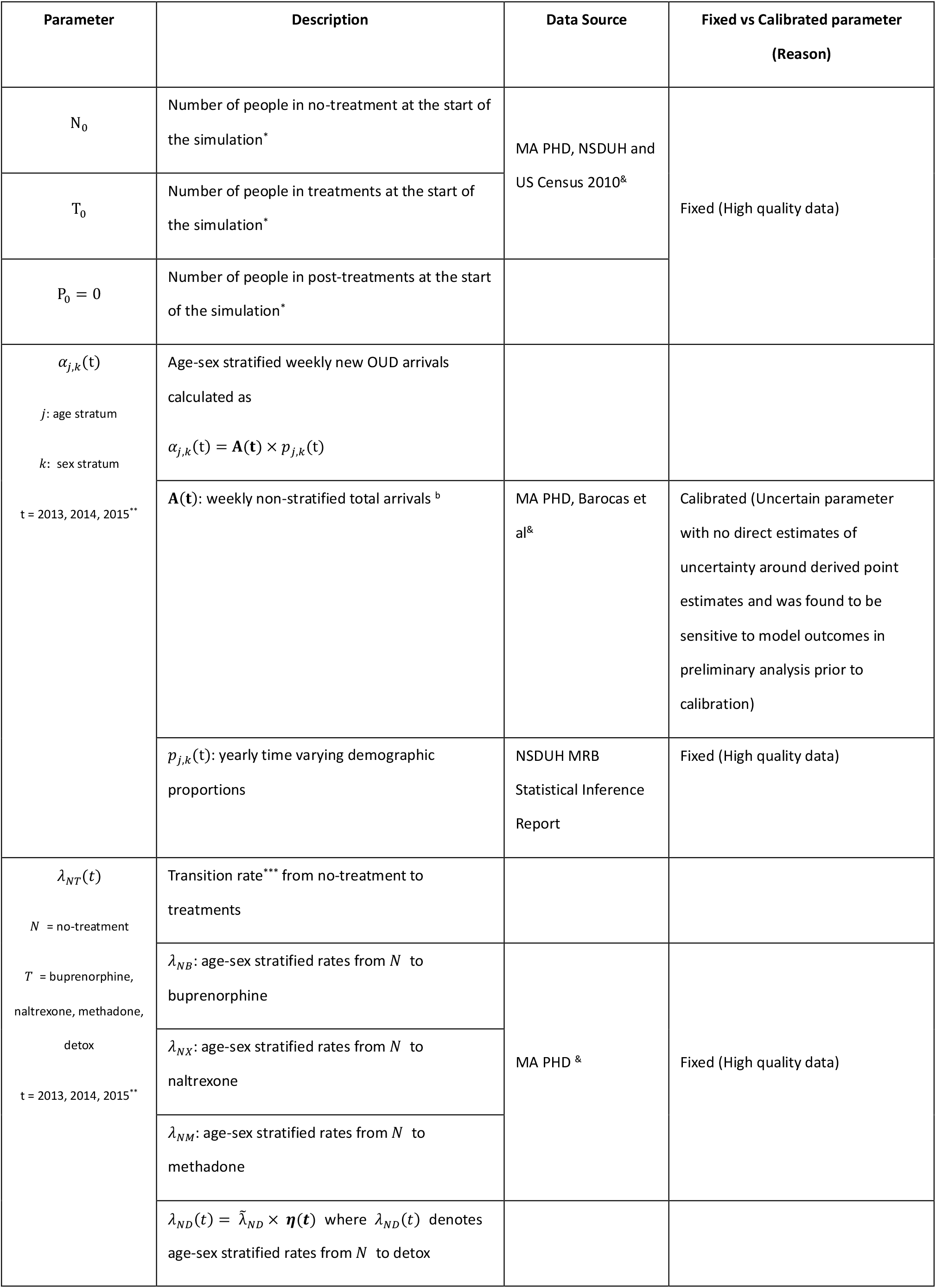
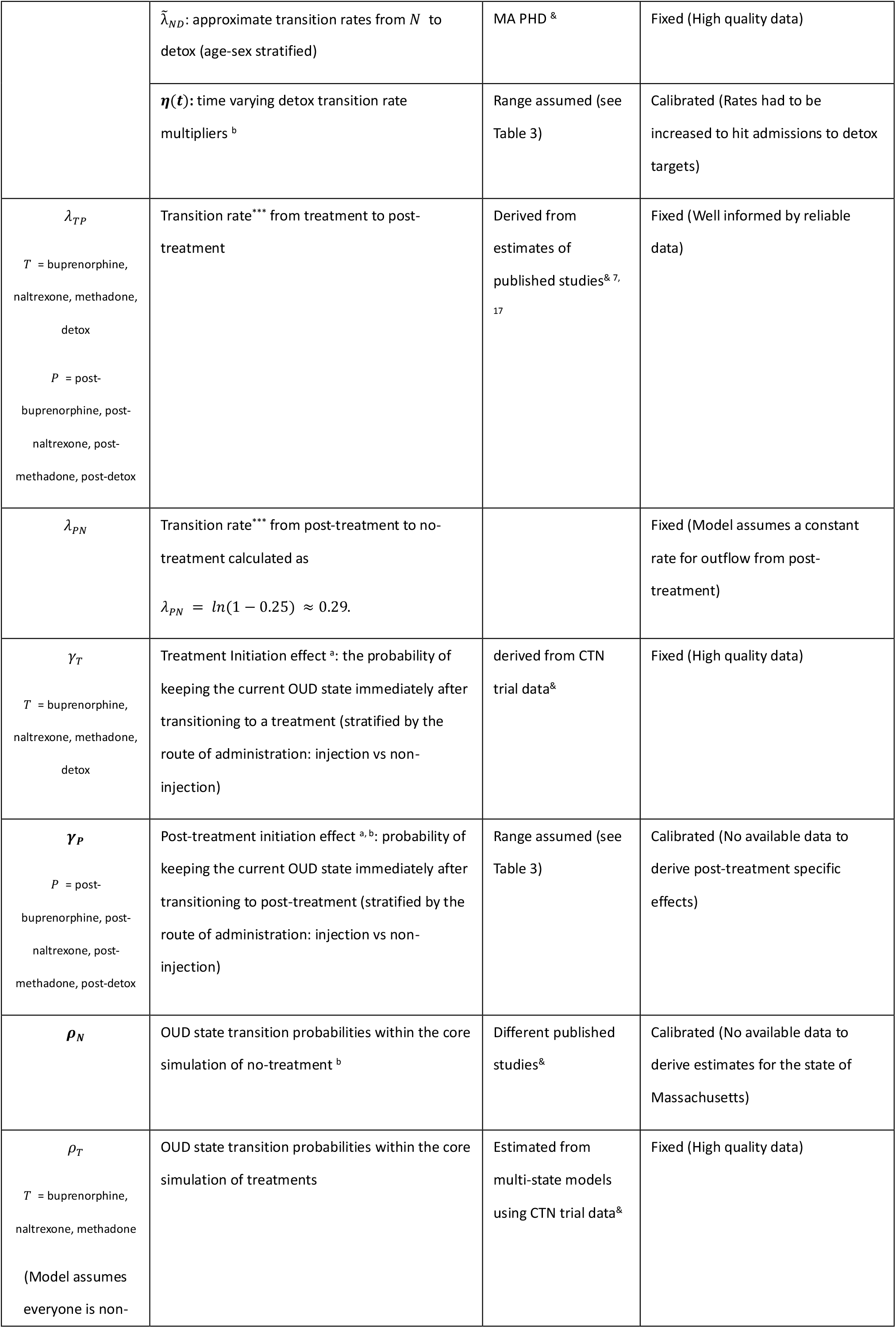
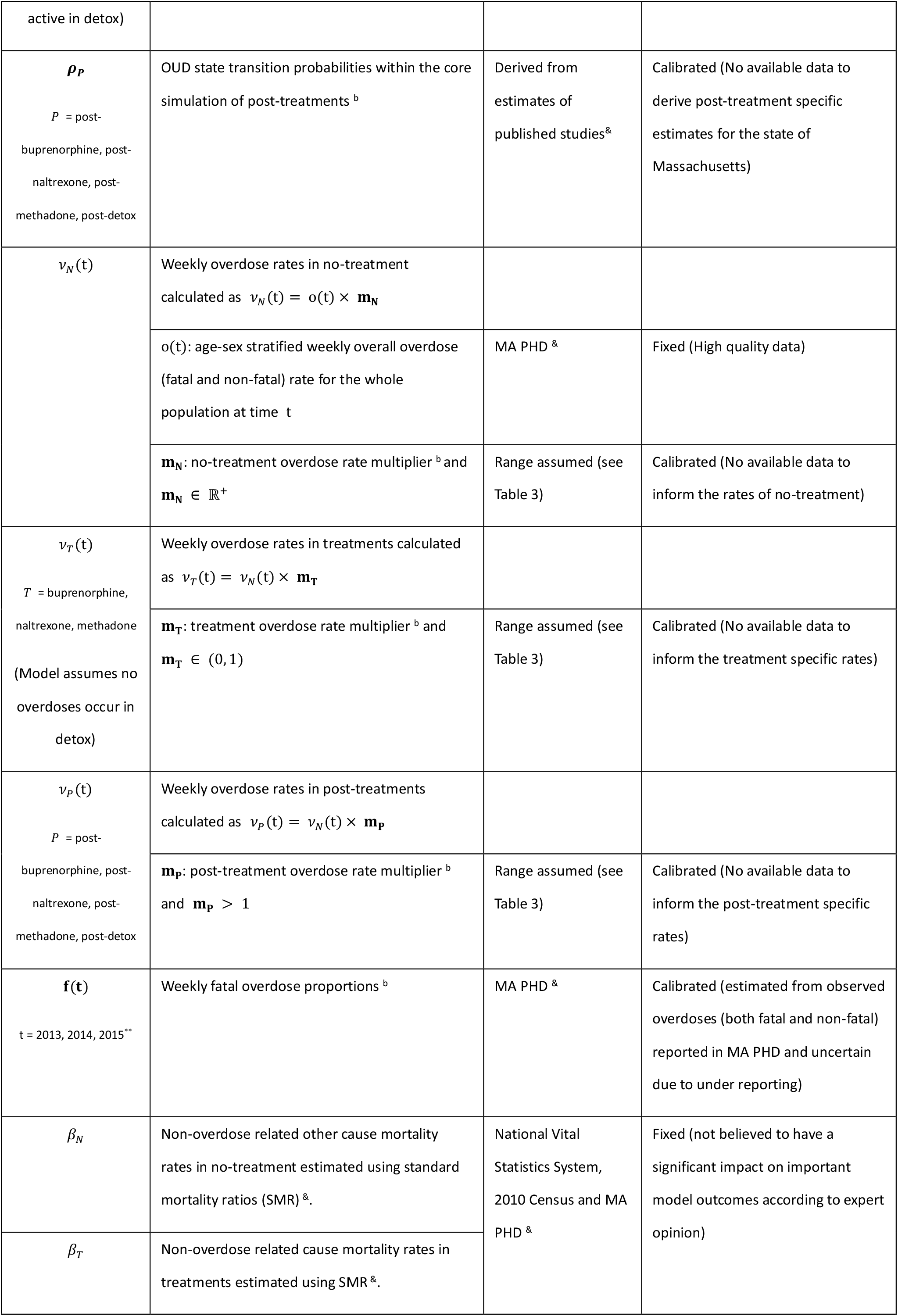
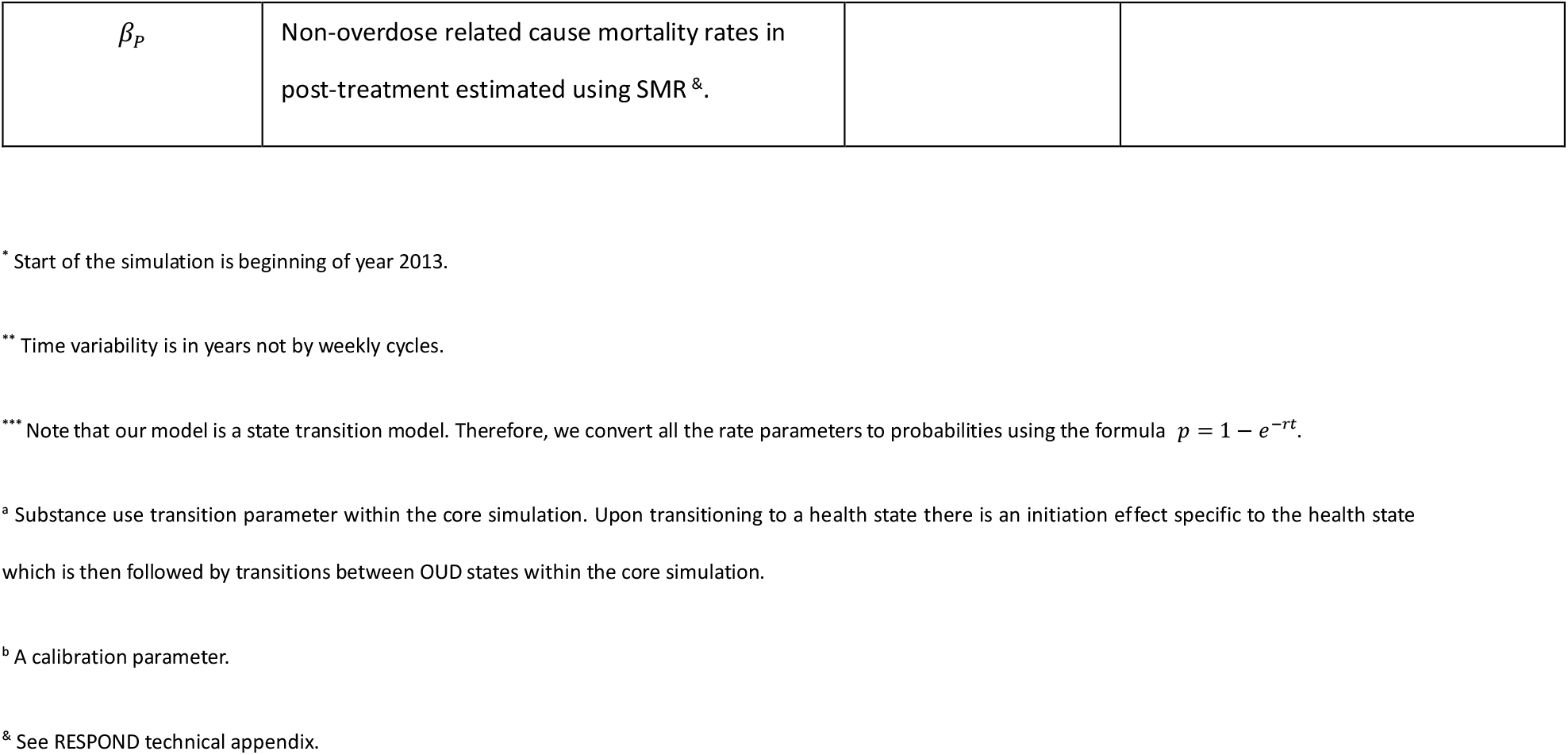
Model parameters.

| Parameter | Description | Data Source | Fixed vs Calibrated parameter (Reason) |
| --- | --- | --- | --- |
| $N_0$ | Number of people in no-treatment at the start of the simulation* | MA PHD, NSDUH and US Census 2010<sup>&</sup> | Fixed (High quality data) |
| $T_0$ | Number of people in treatments at the start of the simulation* | | |
| $P_0 = 0$ | Number of people in post-treatments at the start of the simulation* | | |
| $\alpha_{j,k}(t)$<br>$j$: age stratum<br>$k$: sex stratum<br>$t = 2013, 2014, 2015^{**}$ | Age-sex stratified weekly new OUD arrivals calculated as<br>$\alpha_{j,k}(t) = \mathbf{A}(t) \times p_{j,k}(t)$ | | |
| | $\mathbf{A}(t)$: weekly non-stratified total arrivals <sup>b</sup> | MA PHD, Barocas et al<sup>&</sup> | Calibrated (Uncertain parameter with no direct estimates of uncertainty around derived point estimates and was found to be sensitive to model outcomes in preliminary analysis prior to calibration) |
| | $p_{j,k}(t)$: yearly time varying demographic proportions | NSDUH MRB Statistical Inference Report | Fixed (High quality data) |
| $\lambda_{NT}(t)$<br>$N$ = no-treatment<br>$T$ = buprenorphine, naltrexone, methadone, detox<br>$t = 2013, 2014, 2015^{**}$ | Transition rate*** from no-treatment to treatments | | |
| | $\lambda_{NB}$: age-sex stratified rates from $N$ to buprenorphine | MA PHD <sup>&</sup> | Fixed (High quality data) |
| | $\lambda_{NX}$: age-sex stratified rates from $N$ to naltrexone | | |
| | $\lambda_{NM}$: age-sex stratified rates from $N$ to methadone | | |
| | $\lambda_{ND}(t) = \tilde{\lambda}_{ND} \times \boldsymbol{\eta}(t)$ where $\lambda_{ND}(t)$ denotes age-sex stratified rates from $N$ to detox | | |
| | $\tilde{\lambda}_{ND}$: approximate transition rates from $N$ to detox (age-sex stratified) | MA PHD & | Fixed (High quality data) |
| | $\eta(t)$: time varying detox transition rate multipliers <sup>b</sup> | Range assumed (see Table 3) | Calibrated (Rates had to be increased to hit admissions to detox targets) |
| $\lambda_{TP}$<br>$T$ = buprenorphine, naltrexone, methadone, detox<br>$P$ = post-buprenorphine, post-naltrexone, post-methadone, post-detox | Transition rate*** from treatment to post-treatment | Derived from estimates of published studies<sup>& 7, 17</sup> | Fixed (Well informed by reliable data) |
| $\lambda_{PN}$ | Transition rate*** from post-treatment to no-treatment calculated as<br>$\lambda_{PN} = \ln(1 - 0.25) \approx 0.29$. | | Fixed (Model assumes a constant rate for outflow from post-treatment) |
| $\gamma_T$<br>$T$ = buprenorphine, naltrexone, methadone, detox | Treatment Initiation effect <sup>a</sup>: the probability of keeping the current OUD state immediately after transitioning to a treatment (stratified by the route of administration: injection vs non-injection) | derived from CTN trial data<sup>&</sup> | Fixed (High quality data) |
| $\gamma_P$<br>$P$ = post-buprenorphine, post-naltrexone, post-methadone, post-detox | Post-treatment initiation effect <sup>a,b</sup>: probability of keeping the current OUD state immediately after transitioning to post-treatment (stratified by the route of administration: injection vs non-injection) | Range assumed (see Table 3) | Calibrated (No available data to derive post-treatment specific effects) |
| $\rho_N$ | OUD state transition probabilities within the core simulation of no-treatment <sup>b</sup> | Different published studies<sup>&</sup> | Calibrated (No available data to derive estimates for the state of Massachusetts) |
| $\rho_T$<br>$T$ = buprenorphine, naltrexone, methadone<br>(Model assumes everyone is non- | OUD state transition probabilities within the core simulation of treatments | Estimated from multi-state models using CTN trial data<sup>&</sup> | Fixed (High quality data) |
| active in detox) |  |  |  |
| $\rho_P$<br>$P$ = post-buprenorphine, post-naltrexone, post-methadone, post-detox | OUD state transition probabilities within the core simulation of post-treatments <sup>b</sup> | Derived from estimates of published studies<sup>&</sup> | Calibrated (No available data to derive post-treatment specific estimates for the state of Massachusetts) |
| $v_N(t)$ | Weekly overdose rates in no-treatment calculated as $v_N(t) = o(t) \times m_N$ | | |
| | $o(t)$: age-sex stratified weekly overall overdose (fatal and non-fatal) rate for the whole population at time $t$ | MA PHD <sup>&</sup> | Fixed (High quality data) |
| | $m_N$: no-treatment overdose rate multiplier <sup>b</sup> and $m_N \in \mathbb{R}^+$ | Range assumed (see Table 3) | Calibrated (No available data to inform the rates of no-treatment) |
| $v_T(t)$<br>$T$ = buprenorphine, naltrexone, methadone<br>(Model assumes no overdoses occur in detox) | Weekly overdose rates in treatments calculated as $v_T(t) = v_N(t) \times m_T$ | | |
| | $m_T$: treatment overdose rate multiplier <sup>b</sup> and $m_T \in (0, 1)$ | Range assumed (see Table 3) | Calibrated (No available data to inform the treatment specific rates) |
| $v_P(t)$<br>$P$ = post-buprenorphine, post-naltrexone, post-methadone, post-detox | Weekly overdose rates in post-treatments calculated as $v_P(t) = v_N(t) \times m_P$ | | |
| | $m_P$: post-treatment overdose rate multiplier <sup>b</sup> and $m_P > 1$ | Range assumed (see Table 3) | Calibrated (No available data to inform the post-treatment specific rates) |
| $f(t)$<br>$t = 2013, 2014, 2015^{**}$ | Weekly fatal overdose proportions <sup>b</sup> | MA PHD <sup>&</sup> | Calibrated (estimated from observed overdoses (both fatal and non-fatal) reported in MA PHD and uncertain due to under reporting) |
| $\beta_N$ | Non-overdose related other cause mortality rates in no-treatment estimated using standard mortality ratios (SMR) <sup>&</sup>. | National Vital Statistics System, 2010 Census and MA PHD <sup>&</sup> | Fixed (not believed to have a significant impact on important model outcomes according to expert opinion) |
| $\beta_T$ | Non-overdose related cause mortality rates in treatments estimated using SMR <sup>&</sup>. | | |
| $\beta_p$ | Non-overdose related cause mortality rates in post-treatment estimated using SMR <sup>&</sup>. | | |

<sup>\*</sup> Start of the simulation is beginning of year 2013.
<sup>\*\*</sup> Time variability is in years not by weekly cycles.
<sup>\*\*\*</sup> Note that our model is a state transition model. Therefore, we convert all the rate parameters to probabilities using the formula $p = 1 - e^{-rt}$ .
<sup>a</sup> Substance use transition parameter within the core simulation. Upon transitioning to a health state there is an initiation effect specific to the health state which is then followed by transitions between OUD states within the core simulation.
<sup>b</sup> A calibration parameter.
<sup>&</sup> See RESPOND technical appendix.

### 2.3 Model calibration

The model requires 52 different input parameters, that are stratified by age-sex, by route of administration (injection vs non-injection) and/or time-varying parameters (Table 2, Figure 1). For some of the model parameters we used valid estimates from the literature. We used data from NIDA CTN trials to estimate weekly substance use transition rates in treatment states of MOUDs using a multistate Markov model adjusting for age and sex. Massachusetts state-specific estimates of no-treatment OUD state transition rates are not observable, and we derived these from estimates provided in published studies that used data from different states: New York, California, and Maryland. Similarly, due to the non-availability of data, we assume post-treatments have the same transition rates of no-treatment except for transitions c (NANI → ANI) and g (NAI → AI), for which we assume higher rates compared to no-treatment. Transition rates *c* and *g* in all post-treatment types were derived from estimates published in a study of opioid detoxification patients. Also, there were no data to inform annual new OUD arrival counts. Additionally, parameter estimates of overdose rates and fatal overdose proportions are derived from underreported overdose counts observed from MA PHD. We decided to calibrate unobservable or highly uncertain model parameters, and use fixed values for all the others, for which good data and validate estimates are available.

Here, we calibrate a smaller set of uncertain model parameters informed by less reliable data sources that we found to have an impact on outcomes from preliminary model runs. In Table 2, we summarize the rationale for calibrating (or not) parameters. We calibrate the total number of new OUD arrivals in a week, multipliers on transition rates from no-treatment to detox, multipliers on all types of overdose rates in all heath states, fatal overdose proportions and age-stratified OUD state transition probabilities in no-treatment and post-treatment states (Table 3). The calibration process uses data from 2013 to 2015.

**Table 3.** Calibration parameters.

| Calibrated Parameters | Marginal Distribution | Source |
| --- | --- | --- |
| Weekly non-stratified total arrival counts $\mathbf{A}(t)$<br>$t=2013$<br>$t=2014$<br>$t=2015$ | Uniform (6, 406) *<br>Uniform (7, 307) *<br>Uniform (356, 1356) * | MA PHD * |
| Detox transition rate multiplier $\eta(t)$<br>$t=2013$<br>$t=2014$<br>$t=2015$ | Uniform (1, 2)<br>Uniform (1.25, 2.5)<br>Uniform (1, 2) | Arbitrary<br>selected range<br>learnt through<br>calibration<br>process |
| Post-treatment initiation effects $\gamma_P$ | Uniform (0, 1) | Arbitrary<br>selected range<br>for probabilities |
| OUD state transition probabilities in no-treatment ($\rho_N$) and post-treatments ($\rho_P$) | Beta* | * |
| Overdose rate multipliers<br>$\mathbf{m}_N$<br>$\mathbf{m}_P$<br>$\mathbf{m}_T$<br>$T=\text{Buprenorphine}$<br>$T=\text{Naltrexone}$<br>$T=\text{Methadone}$ | Uniform (0.75, 1.75)<br>Uniform (1, 4)<br>Beta* | 18<br>18<br>18, 19* |
| Weekly fatal overdose proportions $\mathbf{f}(t)$<br>$t=2013$<br>$t=2014$<br>$t=2015$ | Beta* | MA PHD * |

\* See supplementary materials.

#### Calibration targets

Based on their importance for shared decision making of OUD, we selected three sets of calibration targets: yearly counts of total OUDs (total of both alive and dead) in the state, admissions to detox facilities in the state and fatal overdoses from 2013-2015 (Table 1). Since total OUD counts cannot be observed directly from data, we obtained estimates using a capture-recapture analysis.^1^ Both detox admissions and fatal overdose count targets were directly observed from MA PHD. When calibrating to the yearly total OUD, we use the 2015 total OUD counts and constrain our calibration to select parameters that result in an increasing trend in OUD from 2013 to 2015. This is because the 2013 and 2014 total OUD counts from capture-recapture are not directly comparable to the 2015 estimates because they are based on slightly different data. Details of calibration targets are summarized in Table 1 and a detailed explanation of how these targets are derived is provided in the supplementary materials.

#### Calibration approach

We calibrate the model using an empirical approach. More specifically, we use Latin Hypercube Sampling to efficiently search the multidimensional input parameter space. The algorithm accepts proposed parameter values when the respective model outputs are “close” to the observed calibration targets where “closeness” is measured by pre-specified uncertainty ranges around each target. For the 2015 estimated total OUD count target we use 95% confidence intervals (CI) from the capture-recapture analysis, and for detox admissions and fatal overdose counts, we use a range of within 10% of the observed count (Table 1). We accept proposed input samples that result in model outcomes within the uncertainty ranges of every target. As an additional step to accept/reject input samples, we reject proposed samples that resulted in a larger total OUD count for 2013 than 2014, or a larger OUD count for 2014 than 2015. Following are the steps in our empirical calibration algorithm.

**Step 1:** Define plausible ranges and marginal distributions for each calibration parameter (Table 3). These ranges can be obtained from available data sources. If no data is available, ranges can be defined according to expert knowledge or arbitrary and learned through the calibration process.

**Step 2:** From marginal distributions defined in step 1, generate *N* sets of random values from the multidimensional parameter space using Latin hypercube (LH) sampling.

**Step 3:** Run model simulations with proposed input parameter values from step 2 and summarize model outputs to compare with pre-specified calibration targets.

**Step 4:** Accept parameter values resulted in model outcomes within the uncertainty ranges of targets in the following order: 2015 total OUD → admissions to detox → Fatal overdoses. Finally, only accept the parameters that resulted in 2013 total OUD counts < 2014 total OUD counts, and 2014 total OUD counts < 2015 total OUD counts.

**Step 5:** If enough samples were accepted in step 4: Investigate the marginal distributions/ranges of accepted samples to see if the values are concentrated at the lower or upper bounds of their pre-defined ranges. When this happens, shift, or expand the ranges within a plausible set of values and repeat the calibration process until a full distribution is observed.

If no/few sets of parameters accepted:

- Increase *N* and start over the calibration process from step 2.
- If no improvement is observed after increasing *N*, it might be a sign of using invalid plausible ranges for single/multiple parameters or structural issues in the model.
  - Find the problematic model outcome(s) unable to match the corresponding target.
  - Find the most influential parameter(s) for the problematic outcome(s) through a sensitivity analysis or according to expert knowledge.
  - Experiment with expanding or shifting the ranges of the most influential parameter(s) and/or change the model structure and re-run the calibration process.

Our empirical calibration selects multiple sets of input parameter values giving us a way to quantify both parameter uncertainty and the uncertainty around model outcomes. Plausible ranges and marginal distributions used for each calibration parameter are shown in Table 3. For detailed information on how the ranges and distributions were obtained we refer the reader to supplementary material (Appendix S2). To ensure that a sufficient number of input samples were accepted from the calibration process, one needs to repeat the calibration process increasing *N* to achieve a higher grid resolution of the multidimensional parameter space until no significant change is observed in the resulting distributions of the calibration parameters. Given that repeating the calibration process with increasing *N* is computationally demanding, we perform a sensitivity analysis using the set of accepted input parameter samples from the calibration. In this sensitivity analysis, we draw random subsamples from the set of accepted input samples. We investigate if the resulting model outcomes stabilize as we increase the size of the random subsample of inputs, thereby ensuring we have accepted enough input samples.

#### Model validation

We validate the calibrated RESPOND model to evaluate its accuracy in making the projections that are most important for the shared decision-making of OUD. There are multiple types of model validation including face validity, internal validity, cross validity, external validity, and predictive validity.^13^ To validate the RESPOND model, we use three types of validation: internal, external and face validity.

We perform internal validation of the model to verify the accuracy and the consistency of the mathematical equations and programming codes by performing a series of robustness checks and extreme value analysis.

To validate externally, we compare model outcomes to data obtained directly from data sources or published studies that were not used in the model calibration process. Our first validation point is the number of deaths due to competing risks during the calibration period. Other deaths observed from MA PHD are not precise counts, and multiple studies have reported that other deaths are usually 2.4-4.1 times the size of overdose deaths.^14-16^ Therefore, we compare the number of other deaths calculated in the model to the number of other deaths obtained by applying the multipliers in the range 2.4-4.1 on fatal overdose count targets. Therefore, these adjusted other death counts are appropriate targets to investigate the model’s validity externally.

One of the key features of the RESPOND is it produces age-gender stratified model outcomes. Since we only used 2015 total OUD as a calibration target, we validate stratified counts by comparing model outcomes to 2015 age-gender stratified total OUD counts estimated from the capture-recapture analysis (see Figure S4).^1^ Similarly, we validate model outcomes for stratified overdose counts (both all types and fatal overdoses) by comparing to the corresponding age-gender observed counts from MA PHD (see Figure S5).

We also externally validated model-generated year-end all types of overdose counts (fatal and non-fatal overdoses) by comparing to observed overdose counts from MA PHD during 2013-2015. Since all types of overdose counts are usually underreported, our external validation ensured that the calibrated model produces overdose counts higher than the observed counts from MA PHD.

The percentage of active users in different health states of the model is non-observable. However, according to clinical and epidemiological experts of OUD, about 90% in no-treatment should be active users. Similarly, the percentage of active users should be lower in treatments and higher in post-treatment episodes. Therefore, we also assess the face validity of these active OUD percentages coming out from the RESPOND model.

With the proposed empirical approach, we can further investigate and resolve the issues that may arise from the model validation process. The flexibility of the calibration process makes it easy to identify possible reasons for model validation failure. When the calibrated model is unable to hit a validation target, we re-do the calibration excluding the targets one by one from the last to first i.e., in the order: Fatal overdoses, admission to detox, 2015 total OUD. With each calibration target excluded, we investigate whether the model can hit the validation target or not. If the model can hit the validation target after removing a calibration target from the calibration process, we then investigate discrepancies between the accepted input samples to rejected samples from the calibration that included the removed target. Having different (non-overlapping ranges) rejected parameter values from the accepted ones implies that the model requires exploration of these rejected parameter values, suggesting that we should bring more flexibility to the model structure or change the model parametrization such that the model can explore all possible parameter combinations to hit both calibration targets as well as validation targets.

## 3. Results

### 3.1 Model Calibration

We generated 5 million (*N*=5 million) input parameter sets (grid size: 5 million model runs × 102 model parameters) using LH sampling. Of these 5 million sets, 740 sets were accepted in the calibration algorithm. As a result of the way the calibration process is defined, all model outcomes of 2015 total OUD counts, admissions to detox counts and fatal overdoses resulted within their respective uncertainty ranges (Figure 2, Table S1). Even though the model slightly underestimated the total OUD count of 2013-14 estimated from capture-recapture analysis on average, the model was able to produce results that closely match the estimated total OUD count of 2015 (Figure 2(A)).

**Figure 2.**
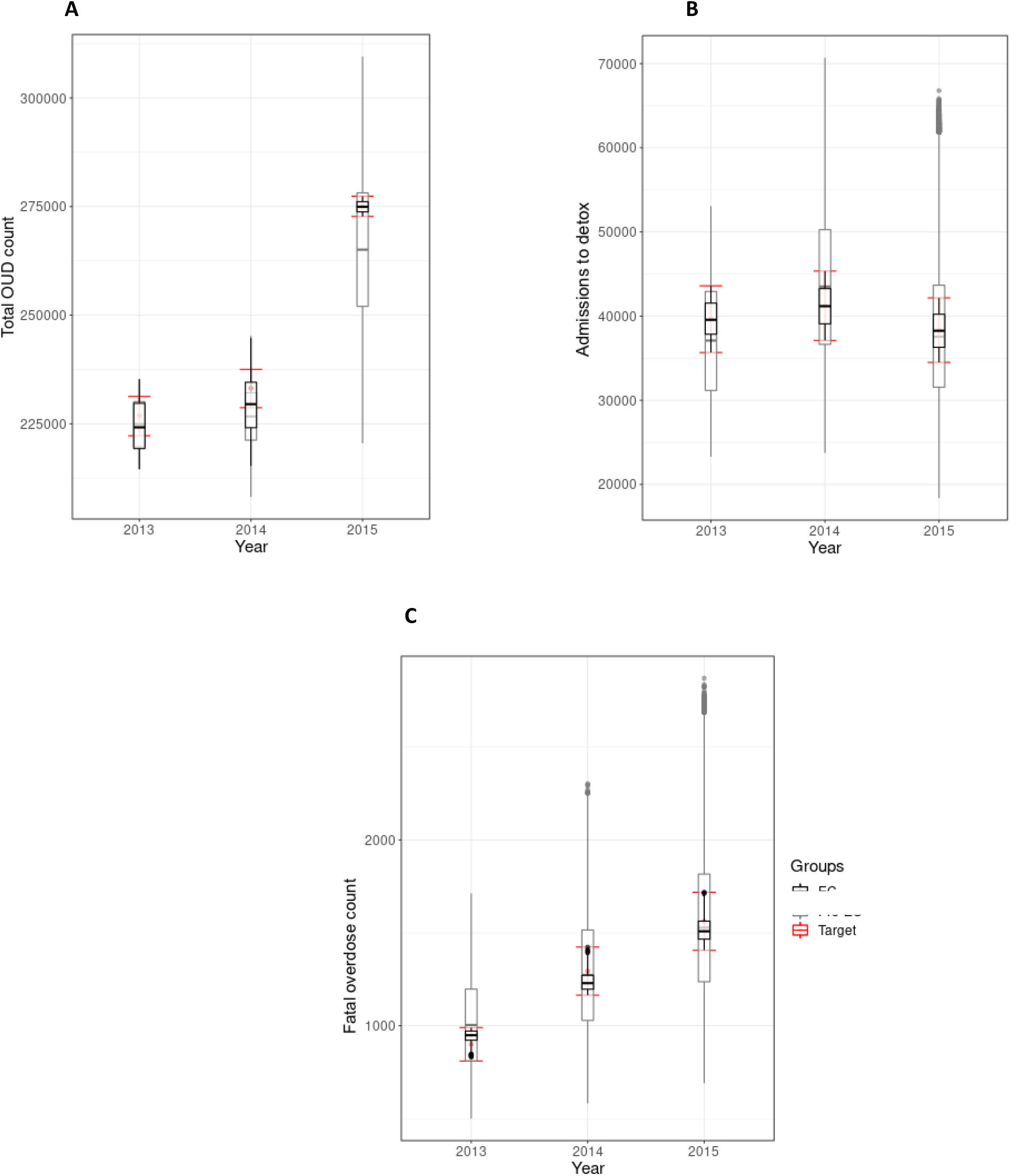
Comparison of calibration targets and ranges to pre and post calibration model outcomes: (A) total OUD counts (B) Admissions to detox counts, and (C) Fatal overdose counts.

We performed a sensitivity analysis to demonstrate that we generated enough LH samples to obtain accurate model outcomes. When we increase the number of samples in the subset above 500, the mean or median values of the model outcomes remain stable (Figure S1), indicating that our final sample size of 740 is sufficient to capture the parameter space.

From the calibration process, we obtained an empirical joint distribution of the calibration parameters (Figure 3). The calibration process substantially changed the marginal distributions of several parameters with non-informative uniform marginal distributions. These are 2014-15 total new OUD arrivals, detox rate transition multipliers, and no-treatment overdose rate multiplier. Prior to calibration, we derived informative marginal distributions from data for overdose rate multipliers in treatments, fatal overdose proportions and OUD transition probabilities in no-treatment and post-treatments prior to the calibration. The calibration process did not change these already informative marginal distributions (see Figure 3 and Figure S2) implying that parameter ranges were already well-defined.

**Figure 3.**
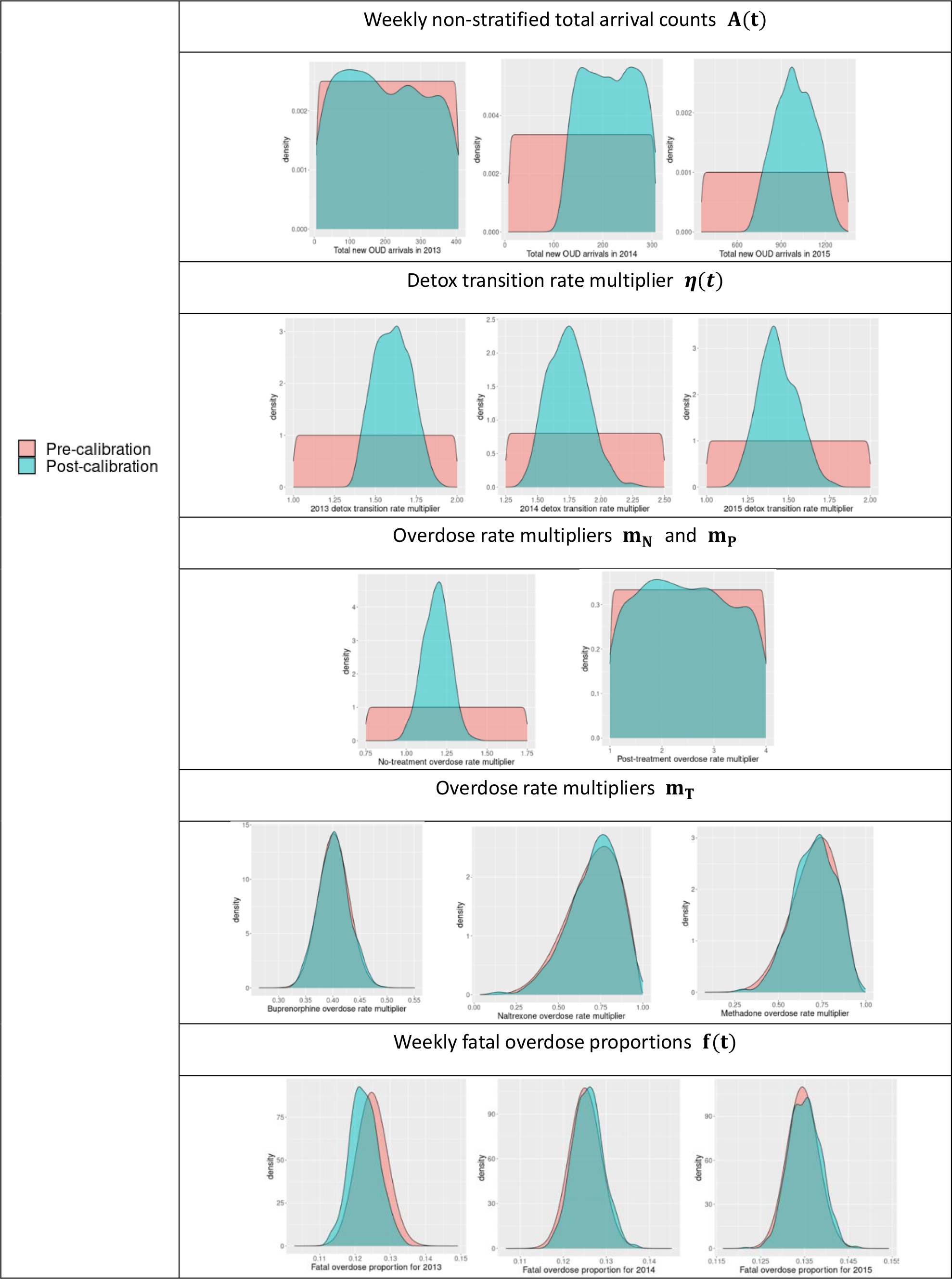
Pre and post calibration comparison of marginal distributions of selected set of calibration parameters. ^**\***^ Marginal distributions of substance use transitions parameters: **γ**_***p***_, ***ρ***_***N***_ and ***ρ***_***p***_ are shown in Figure S2

Calibration failed to identify a more specific marginal distribution than the prespecified uniform distribution for 2013 total new OUD arrivals, post-treatment overdose rate multiplier and initiation effects in post-treatment states. We did not incorporate a direct estimate for total OUD count in 2013 and 2014 in the calibration process likely leading to a dearth of information for new OUD arrivals of these two years although the last filtering step of selecting larger arrivals for 2014 compared to 2013 shifted the prior of 2014 arrivals towards the right end of its range. Similarly, we do not have health state-specific targets to calibrate post-treatment parameters. An overwhelming proportion of OUD was in no-treatment leaving only a small proportion in post-treatments (see Table S2). Correspondingly, overdose counts from no-treatment were significantly higher in no-treatment compared to post-treatments due to a higher percentage of active users (see Table S3). In the current model structure, we calculate yearly fatal overdose counts conditioned on overall all types of overdoses counts, not on overdose counts specific to different health states. As a result, a higher proportion of fatal overdoses occur in no-treatment compared to post-treatment. Since fatal overdose counts are a target, the calibration was dominated by the fatal overdoses of no-treatment leading to an accurate calibration of no-treatment overdose rate multiplier and uninformative calibration of post-treatment overdose rate multiplier.

We did not observe a significant correlation (higher than 0.6 in absolute value) among the parameters except for new OUD arrival counts. We observed a strong negative correlation (r=-0.89) between new OUD arrival counts of 2013 and 2015, with a much weaker correlation between 2014 and 2015 and 2013 and 2014 (r=-0.41 and r-0.005, respectively).

### 3.2 Model Validation

With 740 accepted model input sets, we computed year-end all types of overdose counts for 2013-15 from the model. Figure 4 shows the distribution of all types of overdose model outcomes compared to observed counts from MA PHD. Since overdose counts are usually underreported, we expect the model to result in higher overdose counts compared to what is observed. However, as shown in Figure 4, the model only overestimated the overdose counts in 2013, and for both 2014-15, the model underestimated the overdose counts on average.

**Figure 4.**
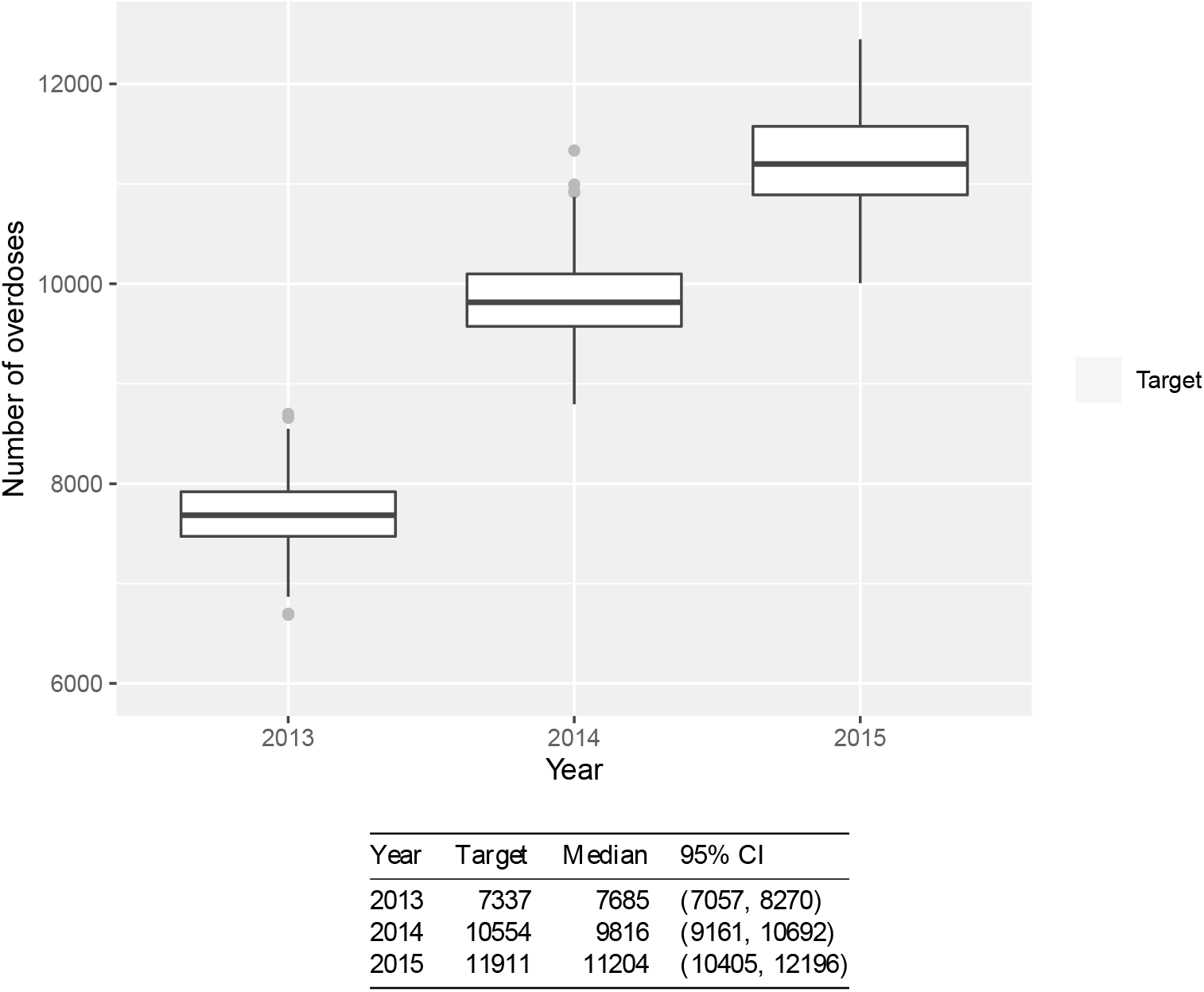
Comparison of year-end all types of overdose counts (both non-fatal and fatal) model outcomes to observed overdose count targets.

To investigate this unexpected result, we excluded the fatal overdose targets (of all three years) from the calibration process and found that the model produced more variable overdose counts that aligned more closely with the observed counts (see Figure S3A). Adding only the 2013 fatal overdose counts target led to an underestimation of the overdose counts of 2014 and 2015 (see Figure S3B). When using only the 2013 fatal overdose target, we found that accepted samples of no-treatment overdose multiplier had non-overlapping ranges to rejected samples (see Figure S3 (C)). Our model uses only one multiplier for no-treatment overdose rates from 2013-15, but this finding suggests that we require time-varying multipliers to produce higher overdose counts for 2014 and 2015.

To validate background death rates, we obtained data from studies reporting overdose death rates and multiplied these by 2.4-4.1 to obtain an estimated external target ranges for background deaths.^14-16^ 2014 and 2015 background death model outcomes were within these target ranges (Figure 5).

**Figure 5.**
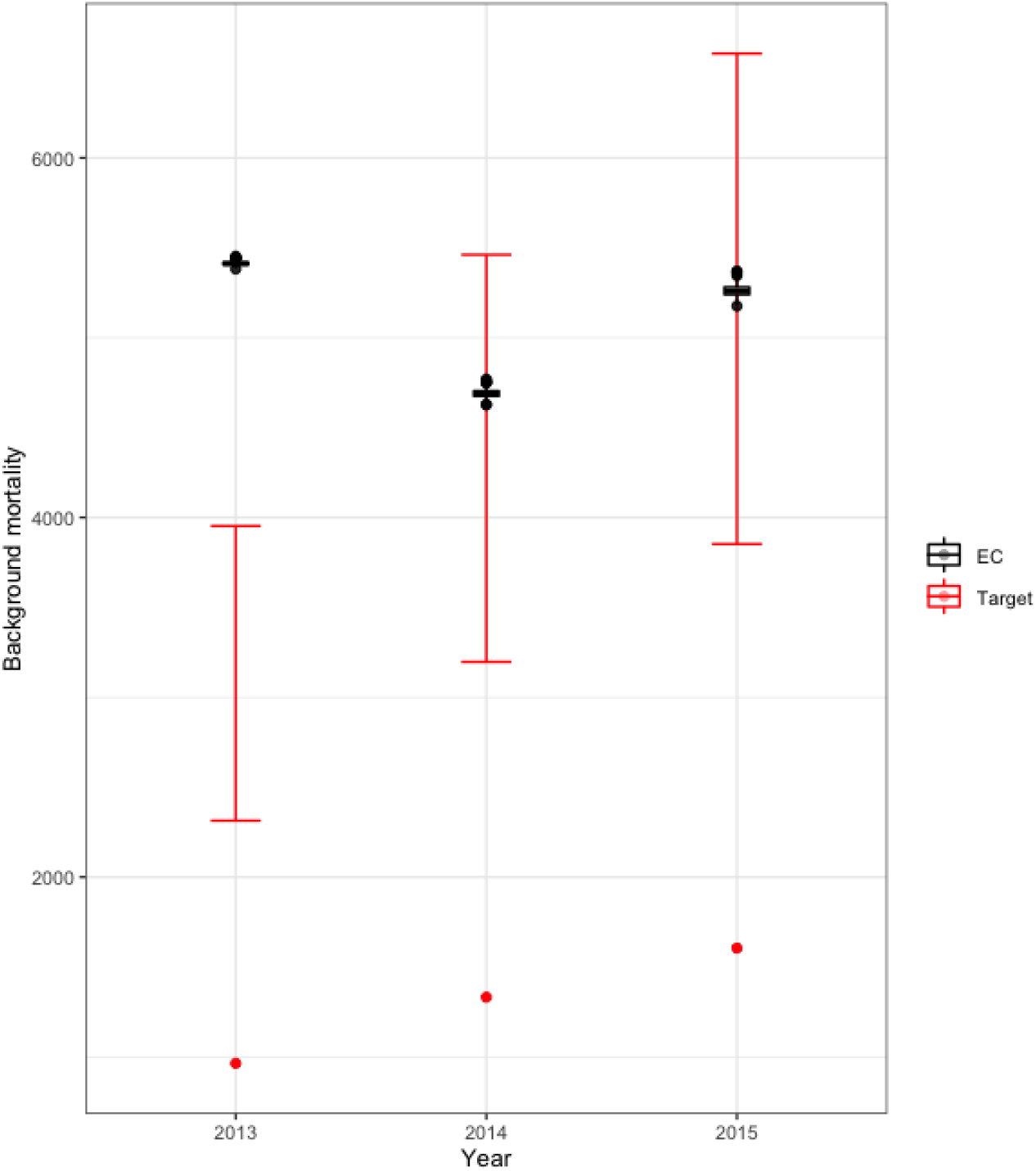
Background/other deaths model outcomes in comparison to target death counts that are corrected to be between 2.4-4.1 times the size of overdose deaths.

From the calibrated model, the median percentage of people who were actively using opioids was 72% (CI: [68%, 77%]) by the end of the year 2013. This median active OUD percentage then declined to 66% (CI: [61%, 75%]) in both 2014 and 2015 (Figure 6A). The median active percentage in no treatment declined from 81% in 2013 to 72% in 2015 but remained stable at 27% and 74% for all treatments and post-treatment episodes in the same time period (Table S4). Even though the model estimated an active percentage in no-treatment lower than expert opinion (90%), the model was able to predict a reasonably higher active OUD percentage (always above 64% from all accepted samples) (Figure 6B). Additionally, the model projects a lower percentage of active OUDs in treatments and a higher percentage in post-treatments meeting face validity criteria.

**Figure 6.**
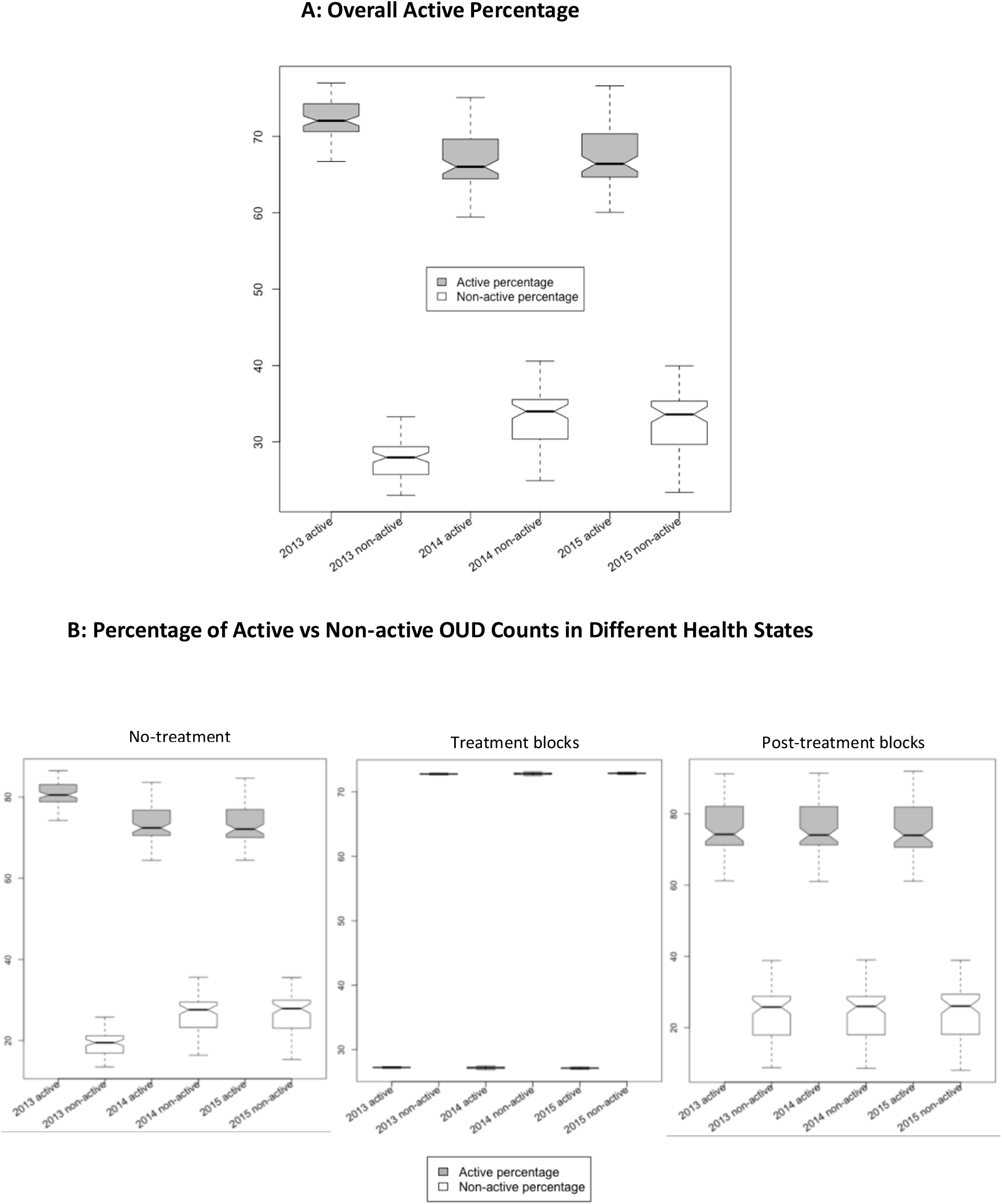
Active vs non-active OUD percentages resulted from the calibrated model. (A) Overall active vs non-active percentages across all health states each year. (B) Yearly percentages of active vs non-active users in different health states

## 4. Discussion

We have provided the details of the calibration and validation process of a dynamic state transition model that simulates the OUD population in Massachusetts. We utilized an empirical calibration approach to fit the model to multiple calibration targets that involved Latin Hypercube sampling to search within the multidimensional input parameter space. The proposed algorithm accepts multiple samples for each parameter providing us a way to quantify the parameter uncertainty simultaneously. From the empirical calibration, we identified precise plausible ranges for model input parameters, and further discovered a set of non-identifiable parameters such as post-treatment parameters due to the model’s structural constraints and lack of available target data. The calibrated model provided a good fit for calibration targets. The model also provided a good fit to a set of validation targets determined to be of the greatest importance such as all types of overdose counts, non-overdose related other death counts and percentages of active users within states. Particularly, the model provided an excellent fit to total OUD counts, fatal overdose counts and background deaths of the year 2015. Therefore, the model outcome of alive OUD counts of 2015 is an accurate prediction of actual alive OUDs in 2015 which can be calculated by subtracting fatal overdose count and background deaths from the total OUD count.

Calibrating complex population models with many parameters is challenging especially when calibration target data is limited. In this paper, we have proposed an empirical calibration approach to calibrate such complex simulation models. In particular, the flexibility of the proposed algorithm allows us to better understand the model structure, identify structural issues and explore underlying relationships between the model parameters. Therefore, utilizing the proposed approach, we gain more insight into the areas where we can simplify the model structurally and/or introduce more target data to improve the model fit.

The proposed empirical calibration algorithm has some limitations. The algorithm can only accept parameter values within the pre-specified range of a marginal distribution and cannot shift the distribution outside of its initial range to fit calibration targets. Therefore, if we create an informative prior distribution for a parameter with a very narrow plausible range it is possible that the calibration will not hit calibration targets. In this case, we suggest expanding the range of the prior and re-running the algorithm. This is distinct from a Bayesian approach, or a mixed type directed search optimization method combined with LH sampling which shifts parameter ranges within a single calibration run by design. However, with such methods, it is usually difficult to achieve convergence for a complex model like RESPOND. The proposed empirical approach informs relevant structural simplifications and produces more informative prior input values to later conduct a more mathematically rigorous calibration exercise.

Further, the proposed empirical approach is a grid-based approach that requires generating a huge amount of LH samples to capture plausible values of input parameter space, as the current filtering approach accepts samples within the uncertainty intervals of each calibration target sequentially. This becomes particularly challenging when extending the calibration time period to incorporate targets of years after 2015 or when calibrating to age-sex stratified targets. Filtering through target ranges one after the other decreases the number of accepted input samples at each filtering step and with many targets we may end up with very few samples or no samples accepted from the calibration process forcing us to increase the size of the LH grid. In a directed search algorithm or a Bayesian calibration, one has to define a single Goodness of Fit measure (GOF) measure like the sum of squared errors or a likelihood to accept/reject samples, and model outcomes from the current calibration can be utilized to define and evaluate a single GOF measure for such a calibration exercise.

The calibration exercise presented here produced a set of non-identifiable parameters, particularly in post-treatments. This is because we lack health state-specific target data and the model structure. The resulting samples of these non-identifiable parameters are still valid as they produce accurate predictions for the model outcomes of interest in this study. However, the model is not well calibrated for applications that primarily focus on post-treatment-specific model outcomes. Further, we did not have good information on prior OUD transition probabilities by age group and our calibration failed to provide age-specific parameter ranges for these probabilities (Figure S2). One implication of this is that we may not need as many age categories since there is not sufficient data to parameterize and calibrate.

One of the limitations in the RESPOND model parametrization is that we assume constant time variability throughout a year for certain parameters such as new OUD arrivals, and overdose rates (both all types and fatal). Even though this framework still allows us to carry out a cost-effective analysis comparing different strategies by projecting future outcomes with the assumption that epidemiological trends observed at the end of the calibration time continue throughout the rest of the simulation, it prevents us from making realistic forecasts from the model such as model projection of OUD population size after 2015. Utilizing data for more years, the parametrization of the RESPOND model can be improved by incorporating time series modeling approaches to model time-dependent parameters.

In the current analysis, even though RESPOND models four types of treatments for OUD, we have incorporated only admission to detox counts as a calibration target. This is because we did not have access to MA PHD to obtain admission counts of other MOUDs at the time of this analysis. Once available, admission to MOUD counts can be used as external targets to validate admission counts resulting from the calibrated model (Figure S6). Incorporating these admission counts as additional targets in our future calibration work should help us to calibrate treatment transition rates in the RESPOND model.

One of the greatest strengths of the RESPOND model is its ability to produce age-gender stratified model outcomes. Depending on the use case of the model, we may require calibrating the model to stratified targets. Current calibration did not consider stratified target counts and as a result, the model failed to produce accurate counts for certain outcomes (see Figure S4 and S5). In a more rigorous Bayesian calibration, we can easily incorporate age-gender stratified targets to derive a log-likelihood enabling the model to produce accurate predictions.

We provided a comprehensive description for the calibration and validation of a complex dynamic population simulation model for OUD. The resulting set of parameter values from the proposed empirical approach will inform the priors of a more comprehensive Bayesian calibration. The calibrated RESPOND model will be employed to improve shared decision-making for OUD.

## Data Availability

All data produced in the present study are available upon reasonable request to the authors

## References

1. Barocas JA, White LF, Wang J, et al. Estimated Prevalence of Opioid Use Disorder in Massachusetts, 2011-2015: A Capture-Recapture Analysis. Am J Public Health 2018: e1–e7. DOI: 10.2105/AJPH.2018.304673.

2. Interim report of the White House Commission on Combating Drug Addiction and the Opioid Crisis. July 31, 2017. Available at: https://www.whitehouse.gov/sites/whitehouse.gov/files/ondcp/commission-interim-report.pdf. Accessed August 24, 2017.

3. Hedegaard H, Miniño AM and Warner M. Drug overdose deaths in the United States, 1999–2018. 2020.

4. Katz, J. Drug Deaths in America Are Rising Faster Than Ever. The New York Times. June 5, 2017. Available at https://www.nytimes.com/interactive/2017/06/05/upshot/opioid-epidemic-drug-overdose-deaths-are-rising-faster-than-ever.html?mcubz=1 Accessed August 25, 2017.

5. Lawrence Scholl PS, Mbabazi Kariisa, Nana Wilson, and Grant Baldwin. Drug and Opioid-Involved Overdose Deaths — United States, 2013–2017. Morbidity and Mortality Weekly Report 2018; 67: 1419–1427.

6. Mattick RP, Breen C, Kimber J, et al. Buprenorphine maintenance versus placebo or methadone maintenance for opioid dependence. Cochrane Database Syst Rev 2014: CD002207. 2014/02/07. DOI: 10.1002/14651858.CD002207.pub4.

7. Morgan JR, Schackman BR, Leff JA, et al. Injectable naltrexone, oral naltrexone, and buprenorphine utilization and discontinuation among individuals treated for opioid use disorder in a United States commercially insured population. J Subst Abuse Treat 2017. DOI: 10.1016/j.jsat.2017.07.001.

8. Barbosa C, Dowd WN and Zarkin G. Economic evaluation of interventions to address opioid misuse: a systematic review of methods used in simulation modeling studies. Value in Health 2020.

9. Chen Q, Larochelle MR, Weaver DT, et al. Prevention of prescription opioid misuse and projected overdose deaths in the United States. JAMA network open 2019; 2: e187621–e187621.

10. Grefenstette JJ, Brown ST, Rosenfeld R, et al. FRED (A Framework for Reconstructing Epidemic Dynamics): an open-source software system for modeling infectious diseases and control strategies using census-based populations. BMC public health 2013; 13: 1–14.

11. Jalali MS, Ewing E, Bannister CB, et al. Data Needs in Opioid Systems Modeling: Challenges and Future Directions. American journal of preventive medicine 2021; 60: e95–e105.

12. Linas BP, Savinkina A, Madushani R, et al. Projected Estimates of Opioid Mortality After Community-Level Interventions. JAMA Netw Open 2021; 4: e2037259. DOI: 10.1001/jamanetworkopen.2020.37259.

13. Eddy DM, Hollingworth W, Caro JJ, et al. Model transparency and validation: a report of the ISPOR-SMDM Modeling Good Research Practices Task Force–7. Medical Decision Making 2012; 32: 733–743.

14. Evans E, Li L, Min J, et al. Mortality among individuals accessing pharmacological treatment for opioid dependence in California, 2006-10. Addiction 2015; 110: 996–1005. 2015/02/04. DOI: 10.1111/add.12863.

15. Hser YI, Mooney LJ, Saxon AJ, et al. High Mortality Among Patients With Opioid Use Disorder in a Large Healthcare System. Journal of addiction medicine 2017; 11: 315–319. 2017/04/21. DOI: 10.1097/ADM.0000000000000312.

16. Larney S, Tran LT, Leung J, et al. All-cause and cause-specific mortality among people using extramedical opioids: a systematic review and meta-analysis. JAMA psychiatry 2020; 77: 493–502.

17. Strain EC, Stitzer ML, Liebson IA, et al. Dose-response effects of methadone in the treatment of opioid dependence. Annals of internal medicine 1993; 119: 23–27.

18. Morgan J, Schackman B, Weinstein Z, et al. Overdose following initiation of naltrexone and buprenorphine medication treatment for opioid use disorder in a United States commercially insured cohort. Drug and Alcohol Dependence 2019; 200: 34–39. DOI: 10.1016/j.drugalcdep.2019.02.031.

19. Sordo L, Barrio G, Bravo MJ, et al. Mortality risk during and after opioid substitution treatment: systematic review and meta-analysis of cohort studies. BMJ 2017; 357: j1550. DOI: 10.1136/bmj.j1550.

