## Supplemental Material for "Empirical Calibration of a Simulation Model of Opioid Use Disorder"

**RESPOND Empirical Calibration Supplementary material**

**Appendix S1.** RESPOND Model Structure and Parameter Estimation

**Appendix S2.** Derivation of Marginal Distributions for Calibration Parameters

**Appendix S3.** Calibration Targets

**Figure S1.** Results from the Sensitivity Analysis of Accepted Model Outcomes

**Figure S2.** Pre-calibration Marginal Distributions vs Post-calibration Marginal Distributions of Substance Use Transitions Parameters

**Figure S3.** Accepted vs Rejected Samples of No-treatment Overdose Multiplier

**Figure S4.** Model External Validity for Age-gender Stratified Total OUD Counts

**Figure S5.** Model External Validity for Age-gender Stratified Overdose Counts

**Figure S6.** Model Validity for Admissions to Treatment

**Table S1.** Model Outcomes vs Calibration Targets

**Table S2.** Alive OUD Percentages

**Table S3.** Overall Active OUD Percentages Distributed among Health States

**Table S4.** Active OUD Percentages

**References**

#### Appendix S1. RESPOND Model Structure and Parameter Estimation

For a detailed overview of the RESPOND model and estimation of base case parameter values, we refer the reader to the comprehensive documentation “the RESPOND user guide” supplemental located at “https://www.syndemicslab.org/respond” For easy navigation, Table S1.1 summarize the location where reader can find detail information on model parameters from the RESPOND user guide. Further, we provide additional details on derivations of marginal distributions of calibration parameters and derivation of calibration targets particular to the empirical calibration in following sections Appendix S2 and Appendix S3 respectively.

Table S1.1. Location information for the details of model parameters described in RESPOND user guide

| Parameter | Description | Location in RESPOND user guide |
|---|---|---|
| $\mathrm{N}_0$ | Number of people in no-treatment at the start of the simulation[*] | Section C.2.1.1 |
| $\mathrm{T}_0$ | Number of people in treatments at the start of the simulation[*] | |
| $\mathrm{P}_0 = 0$ | Number of people in post-treatments at the start of the simulation[*] | |
| $\alpha_{j,k}(\mathrm{t})$<br>$j$: age stratum<br>$k$: sex stratum<br>t = 2013, 2014, 2015[**] | Age-sex stratified weekly new OUD arrivals calculated as<br>$\alpha_{j,k}(\mathrm{t}) = \mathbf{A}(\mathbf{t}) \times p_{j,k}(\mathrm{t})$ | |
| | $\mathbf{A}(\mathbf{t})$: weekly non-stratified total arrivals [b] | Section C.2.1.3 (Table 3) |
| | $p_{j,k}(\mathrm{t})$: yearly time varying demographic proportions | Section C.2.1.3 (Table 2) |
| $\lambda_{NT}(t)$<br>$N$ = no-treatment<br>$T$ = buprenorphine, naltrexone, methadone, detox<br>t = 2013, 2014, 2015[**] | Transition rate[***] from no-treatment to treatments | Section C.2.3.1 (Table 5 presents the converted weekly probabilities calculated from transition rates) Note that the probabilities provided under the “Detox” column in Table 5 are pre-calibration approximate probabilities. |
| | $\lambda_{NB}$: age-sex stratified rates from $N$ to buprenorphine | |
| | $\lambda_{NX}$: age-sex stratified rates from $N$ to naltrexone | |
| | $\lambda_{NM}$: age-sex stratified rates from $N$ to methadone | |
| | $\lambda_{ND}(t) = \tilde{\lambda}_{ND} \times \boldsymbol{\eta}(\boldsymbol{t})$ where $\lambda_{ND}(t)$ denotes age-sex stratified rates from $N$ to detox | |
| | $\tilde{\lambda}_{ND}$: approximate transition rates from $N$ to detox (age-sex stratified) | |
| | $\boldsymbol{\eta}(\boldsymbol{t})$**:** time varying detox transition rate multipliers [b] | |
| $\lambda_{TP}$<br>$T$ = buprenorphine, naltrexone, methadone, detox<br>$P$ = post-buprenorphine, post-naltrexone, post-methadone, post-detox | Transition rate[***] from treatment to post-treatment | Section C.2.3.4 (Table 8) |
| $\lambda_{PN}$ | Transition rate[***] from post-treatment to no-treatment calculated as<br>$\lambda_{PN} = ln(1 - 0.25) \approx 0.29.$ | |
| $\gamma_T$<br>$T$ = buprenorphine, naltrexone, methadone, detox | Treatment Initiation effect [a]: the probability of keeping the current OUD state immediately after transitioning to a treatment (stratified by the route of administration: injection vs non-injection) | Section C.2.3.2 (Table 6) |
| $\boldsymbol{\gamma_P}$<br>$P$ = post-buprenorphine, post-naltrexone, post-methadone, post-detox | Post-treatment initiation effect [a, b]: probability of keeping the current OUD state immediately after transitioning to post-treatment (stratified by the route of administration: injection vs non-injection) | Section C.2.3.2 (Table 6) |
| $\boldsymbol{\rho_N}$ | OUD state transition probabilities within the core simulation of no-treatment [b] | Section C.2.2 (Table 4) |
| $\rho_T$<br>$T$ = buprenorphine, naltrexone, methadone<br>(Model assumes everyone is non-active in detox) | OUD state transition probabilities within the core simulation of treatments | Section C.2.3.3 (Table 7) |
| $\boldsymbol{\rho_P}$<br>$P$ = post-buprenorphine, post-naltrexone, post-methadone, post-detox | OUD state transition probabilities within the core simulation of post-treatments [b] | Section C.2.2 (Table 4) |
| $\nu_N(\mathrm{t})$ | Weekly overdose rates in no-treatment calculated as $\nu_N(\mathrm{t}) = \mathrm{o}(\mathrm{t}) \times \mathbf{m_N}$ | Section C.2.4.2 |

| | | |
|---|---|---|
| | $o(t)$: age-sex stratified weekly overall overdose (fatal and non-fatal) rate for the whole population at time t | Section C.2.4.1 (Table 10) |
| | $\mathbf{m_N}$: no-treatment overdose rate multiplier [b] and $\mathbf{m_N} \in \mathbb{R}^+$ | |
| $\nu_T(t)$<br>$T$ = buprenorphine, naltrexone, methadone<br>(Model assumes no overdoses occur in detox) | Weekly overdose rates in treatments calculated as $\nu_T(t) = \nu_N(t) \times \mathbf{m_T}$ | Section C.2.4.3 |
| | $\mathbf{m_T}$: treatment overdose rate multiplier [b] and $\mathbf{m_T} \in (0, 1)$ | Section C.2.4 (Table 11) |
| $\nu_P(t)$<br>$P$ = post-buprenorphine, post-naltrexone, post-methadone, post-detox | Weekly overdose rates in post-treatments calculated as $\nu_P(t) = \nu_N(t) \times \mathbf{m_P}$ | Section C.2.4.4 |
| | $\mathbf{m_P}$: post-treatment overdose rate multiplier [b] and $\mathbf{m_P} > 1$ | Section C.2.4 (Table 11) |
| $\mathbf{f(t)}$<br>t = 2013, 2014, 2015[**] | Weekly fatal overdose proportions [b] | Section C.2.5.1 (Table 12) |
| $\beta_N$ | Non-overdose related other cause mortality rates in no-treatment estimated using standard mortality ratios (SMR). | Section C.2.5.2 |
| $\beta_T$ | Non-overdose related cause mortality rates in treatments estimated using SMR. | |
| $\beta_P$ | Non-overdose related cause mortality rates in post-treatment estimated using SMR. | |

[*] Start of the simulation is beginning of year 2013.

[**] Time variability is in years not by weekly cycles.

[***] Note that our model is a state transition model. Therefore, we convert all the rate parameters to probabilities using the formula $p = 1 - e^{-rt}$.

[a] Substance use transition parameter within the core simulation. Upon transitioning to a health state there is an initiation effect specific to the health state which is then followed by transitions between OUD states within the core simulation.

[b] A calibration parameter.

**Appendix S2. Derivation of Marginal Distributions for Calibration Parameters**

In this section, we describe how we derive marginal distributions for calibration parameters from available data sources. Note that for some calibration parameters (i.e., parameters: $\boldsymbol{\eta}(\boldsymbol{t})$, $\boldsymbol{\gamma_P}$, $\mathbf{m_N}$, and $\mathbf{m_P}$ in Table S1.1) we choose arbitrary ranges with uniform distributions due to no available data to inform the uncertainty, and those parameters are not described here.

**S2.1 Weekly non-stratified total arrival counts $\mathbf{A(t), t = 2013, 2014, 2015}$**

We calculated point estimates for $\mathbf{A(t)}$ from yearly arrival counts (also called entering cohort counts) $N_{enter,t}$ (See Table 3 in RESPOND user guide) as follows.

$$\mathbf{A(t)} = \frac{N_{enter,t}}{52}$$

We used within $x\%$ of these point estimates as the uncertainty range of each uniform marginal distribution. Appropriate value for $x$ were obtained through calibration process by expanding the ranges such that calibration resulted in accepted parameter values.

**S2.2 OUD state transition probabilities in no-treatment ($\boldsymbol{\rho_N}$) and post-treatments ($\boldsymbol{\rho_P}$)**

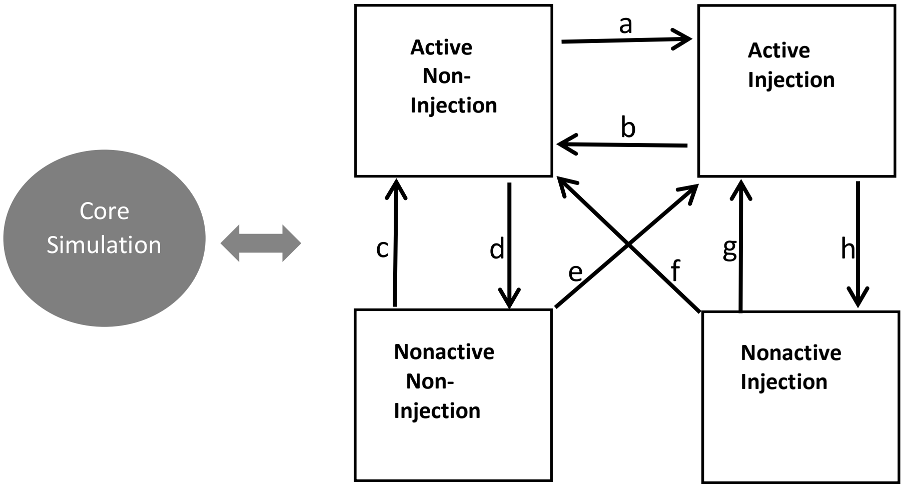


OUD state transition probabilities (denoted by $a, b, c, d, e, f, g$ and $h$) in the core simulation of RESPOND have set of constraints imposed on them. Following we list those constraints.

- $b < a$
- $e, f < g, c$ (Here $e$ and $f$ are independent from each other and similarly $c$ and $g$ are independent from each other. In other words, it is similar to $max(e, f) < min(c, g)$ )
- $d$ and $h$ are independent from each other
- $g > h$ and $c > d$

For each independent transition probability, we fit beta distribution marginals using bootstrap sample draws. To draw bootstrap samples, we used the same distributions that were used when calculating confidence intervals (CI) in the respective literature from where we obtained base case point estimates for OUD transition probabilities (See Table 4 in RESPOND user guide). For dependent transitions with constraints (for example consider $b < a$), we derived constrained marginal distributions as explained below.
Let $a \sim beta(\alpha_1, \theta_1)$, and the risk ratio for $b$ vs $a$ be $rr_{b,a} \sim beta(\alpha_2, \theta_2)$.
Then, $b \sim a \times rr_{b,a}$.
In other words, we fitted a beta distribution for the probability $a$ using bootstrap samples and similarly, we generated bootstrap samples for $rr_{b,a}$ and fitted a beta distribution. The constrained draws for $b$ were then obtained by multiplying the draws of these fitted two beta distributions.

#### S2.3 Treatment overdose rate multipliers $\mathbf{m_T}$, $\mathbf{T} = \boldsymbol{Buprenorphine, Naltrexone, Methadon}$

Overdose rates in treatments are calculated by applying a multiplier $\mathbf{m_T}$ on no-treatment overdose rates. Since we assume in RESPOND that overdose rates in treatments are lower compared no-treatment, $\mathbf{m_T} \in (0, 1)$. Therefore, we fitted beta distributions for $\mathbf{m_T}$ of all three treatment types. All three beta distributions were fitted on bootstrapped samples.

We obtained point estimates for multipliers of buprenorphine and naltrexone from a published study (see Table 10 in RESPOND user guide).[2] These point estimates are hazard ratios from a Cox hazards model. Therefore, we generated bootstrap samples for $\mathbf{m_{Buprenorphine}}$, and $\mathbf{m_{Naltrexone}}$ by taking $log(hazard\ ratio)$ to be normally distributed, and fitted beta distributions on these bootstrapped samples.

Point estimate for methadone overdose rate multiplier $\mathbf{m_{Methadone}}$ is calculated as $\mathbf{m_{Methadone}} = \mathbf{m_{Buprenorphine}} \times \frac{r_\mathrm{m}}{r_\mathrm{b}}$ (see Table 10 in RESPOND user guide).
Here $r_\mathrm{m}$, and $r_\mathrm{b}$ are rate for methadone and buprenorphine respectively.
Estimates for $r_\mathrm{m}$, and $r_\mathrm{b}$ were obtained from a published study.[3] To draw bootstrap samples for $r_\mathrm{m}$, and $r_\mathrm{b}$, we used the distributional assumptions that were used when calculating confidence intervals (CI) in the respective literature.

#### S2.4 Weekly fatal overdose proportions $\mathbf{f(t)}$, $\mathbf{t = 2013, 2014, 2015}$

Since $\mathbf{f(t)}$ are bounded by 0 and 1, we fitted beta marginal distributions. For each $\mathbf{f(t)}$, we first calculated the mean $\mu$ and the standard deviation $\sigma$ from empirical CI (see Table 12 in RESPOND user guide) and then derived the corresponding parameters $\alpha$ and $\beta$ of beta distribution as follows.

$$\alpha = (\frac{1-\mu}{\sigma^2} - \frac{1}{\mu}) \times \mu^2$$

$$\beta = \alpha \left(\frac{1}{\mu} - 1\right)$$

#### Appendix S3. Calibration Targets

##### S3.1 Yearly Total OUD counts

For each year, the total size of population with OUD was estimated using a capture-recapture analysis. We used seven datasets from MA Public Health Datawarehouse as sampling units in a capture-recapture process, including APCD; Bureau of Substance Addiction Services (BSAS); Hospital, ED, and Outpatient Observation discharges (Case Mix); Death certificates (Death); Birth certificates (Birth); Emergency Medical Service Incident Data (MATRIS); and Prescription Monitoring Program (PMP). Year 2013 has incomplete data in MATRIS as it was under the data collection phase, making a limitation on using the estimate for that year as a target. Year 2015 is the most recent year that had data available to us, it was assumed to have collected most complete data than 2013 and 2014. Variables were selected within each of datasets to identify the target population each year. We developed contingency tables that were stratified by age and sex to estimate the total number of known individuals with OUD. There are three groups of age: 11-24, 25-45, and 45 and above. Target population with unknown status of age or sex were removed from the analysis. We then developed stratified log-linear models to estimate the unobserved OUD. Models allowed interaction terms to account for the dependencies between data sources. We used AIC and covariate significance to select the final models for each stratum. A state-level estimate was obtained by summing the strata-level estimates. We computed 95% confidence intervals (CI) of aggregated estimates using parametric bootstrapping.

##### S3.2 Yearly Accumulated Detox Admissions

We used Acute Treatment Service (ATS)/Detox information extracted from Bureau of Substance Addiction Services (BSAS) database to calculate the cases of admission to detox services. For each year between 2013 and 2015, we first used abovementioned seven data sources, along with the same variables, to label an OUD status to subjects appeared to ATS/Detox program in BSAS. Among the observed subjects who were identified with OUD, the number of admission cases stratified by age and sex were counted as the target data. Target population with unknown status of age or sex were removed from the calculation.

##### S3. 3 Yearly Accumulated Fatal Overdose Counts

We used Death certificates database to calculate the death that involved opioid overdose. We assumed that all the death with opioid overdose involved were people with OUD and no person dead due to OUD was incorrectly missed. We first used abovementioned seven data sources, along with the same variables, to label an OUD status to subjects appeared in the Death Certificates. Among the observed subjects who were identified with OUD, the number of death due to opioid overdose were stratified by age and sex and counted as the target data. Target population with unknown status of age or sex were removed from the calculation.

**Figure S1. Results from the Sensitivity Analysis of Accepted Model Outcomes**

**A**

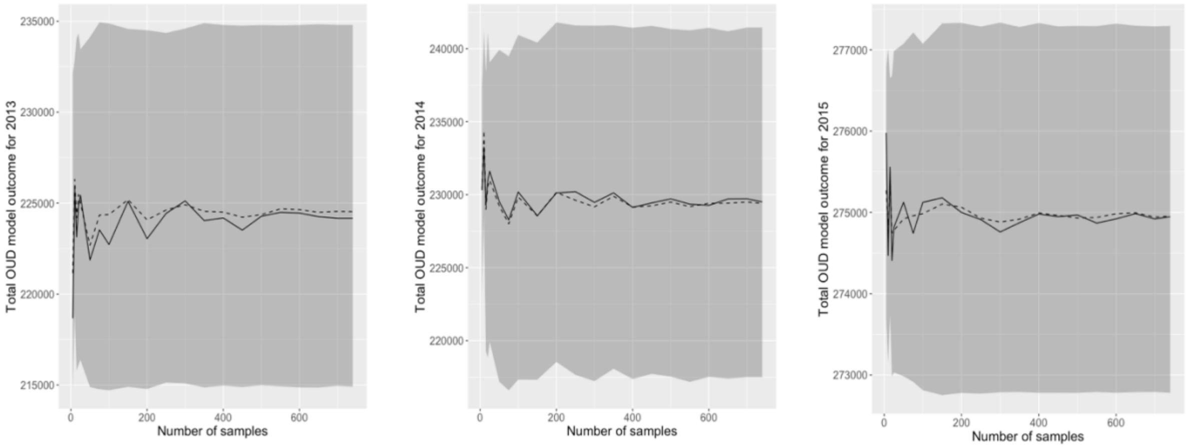


**B**

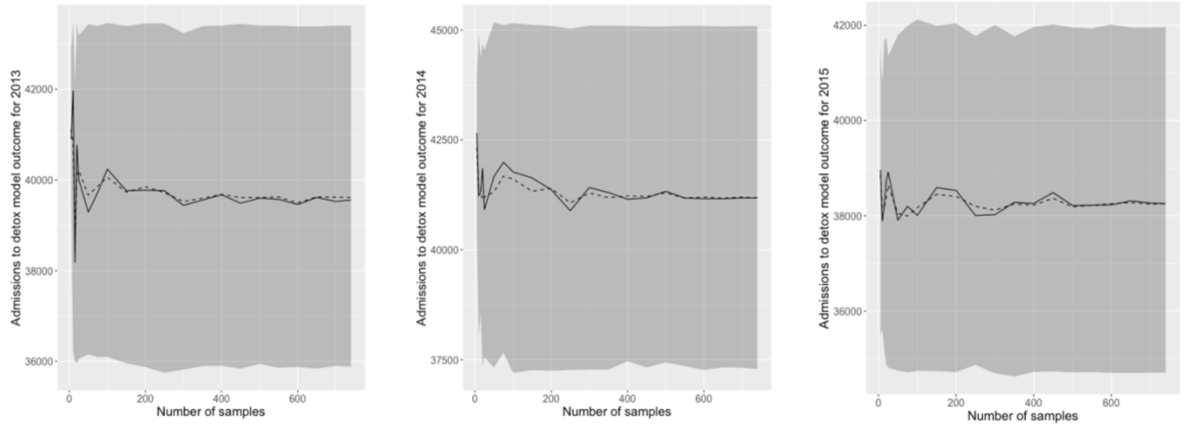


**C**

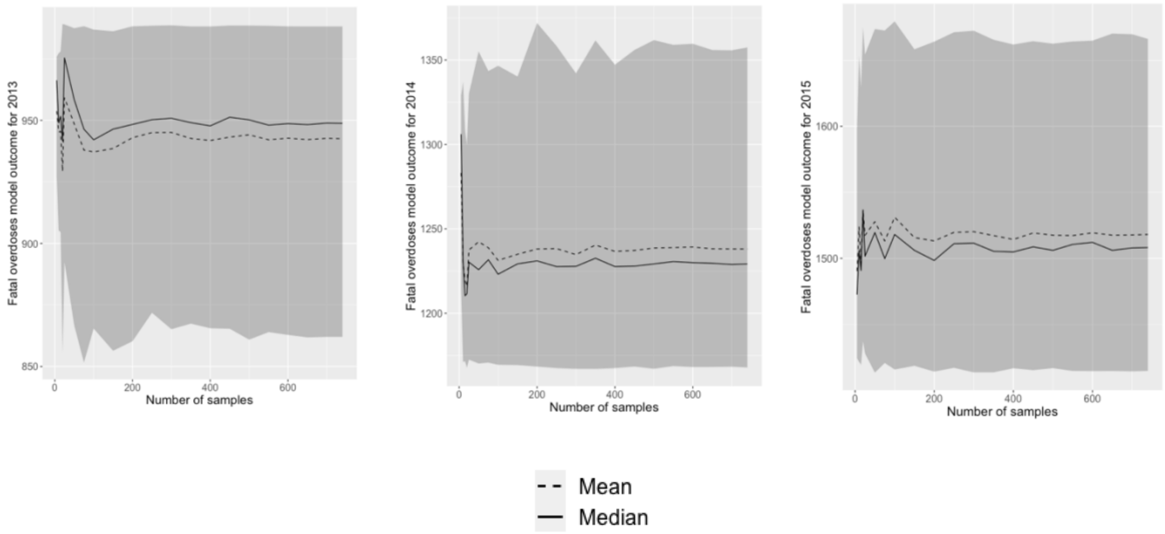


Figure S1 shows the variability of model outcomes as we increase the size of the accepted model inputs sample.

**Figure S2: Pre-calibration Marginal Distributions vs Post-calibration Marginal Distributions of Substance Use Transitions Parameters**

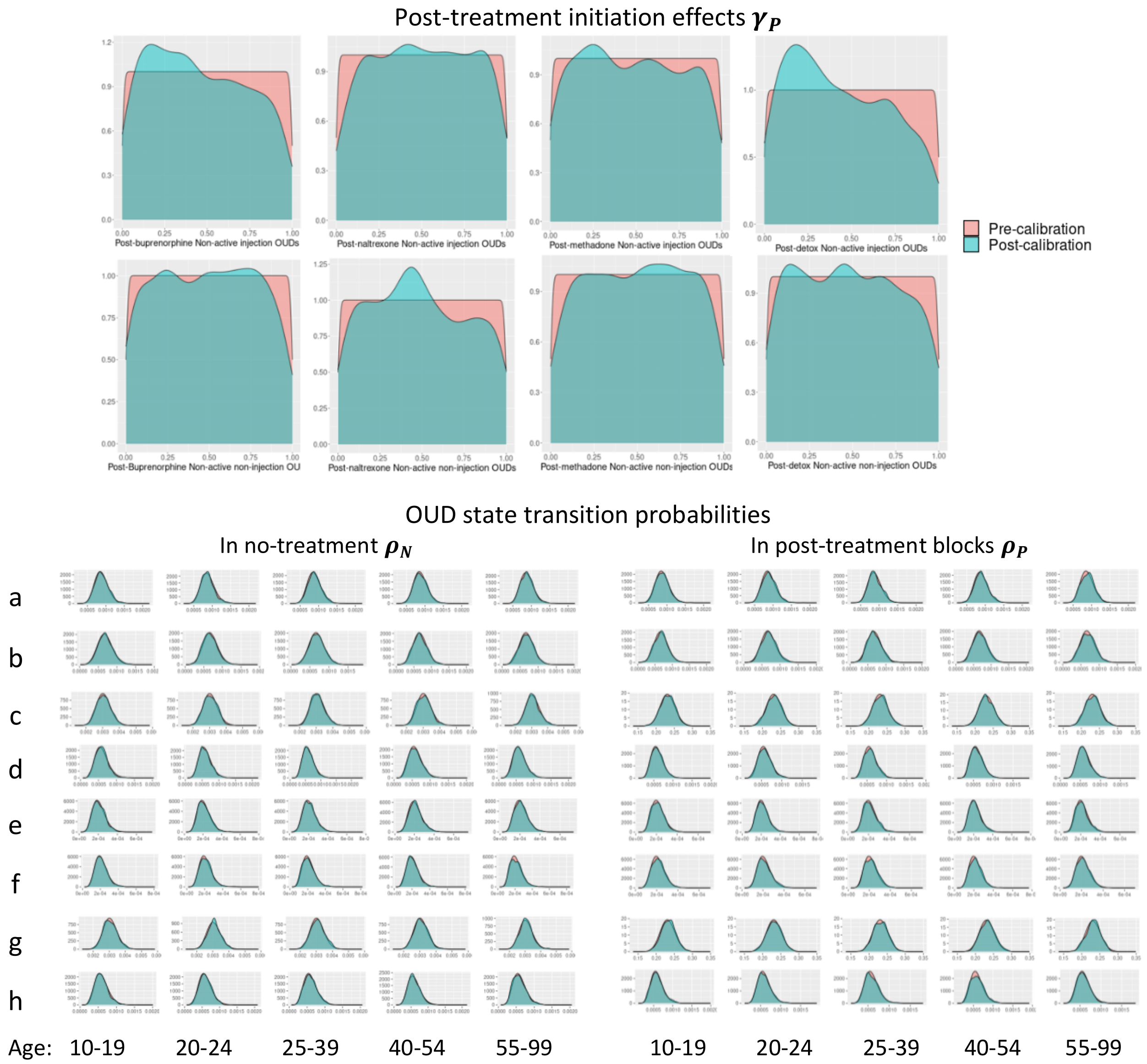

#### Figure S3: Accepted vs Rejected Samples of No-treatment Overdose Multiplier

**A**

Calibrating without fatal overdose counts targets

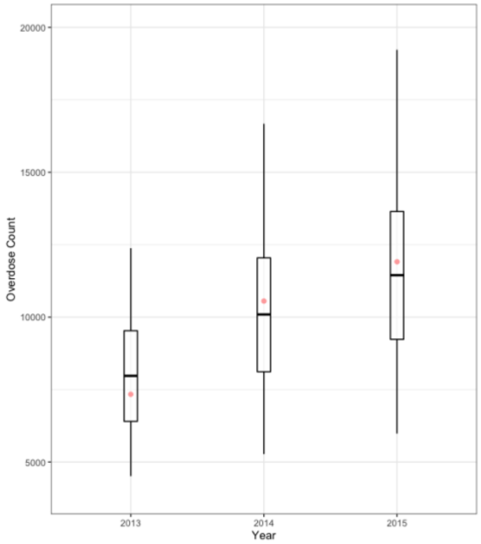


**B**

After adding fatal overdose counts of 2013 target

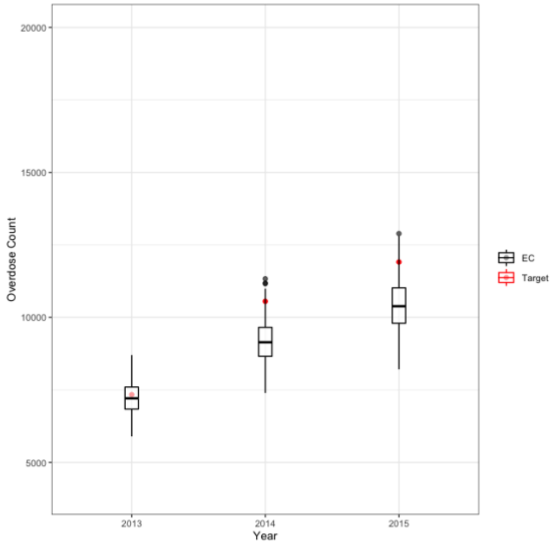


**C**

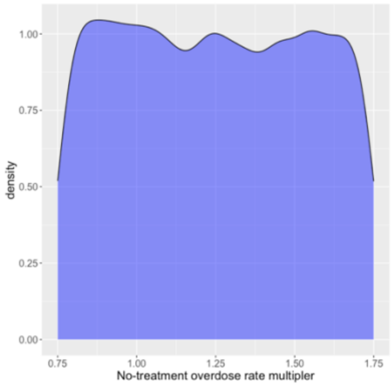


Accepted input samples when calibrating to total OUD counts and admissions to detox excluding fatal overdose counts targets

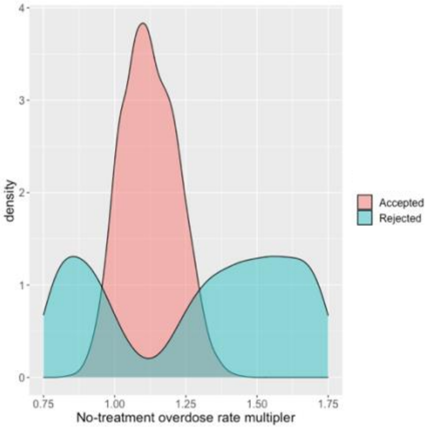


Accepted vs Rejected input samples further calibrating to fatal overdose count of 2013

Figure S3 shows the behavior of all types of overdose counts model outcomes when excluding and including a calibration target. (A) Compare the observed counts of year-end all types of overdoses to the counts resulted from the model when we exclude fatal overdose counts targets (of all three years) from the calibration process. (B) Compare the observed counts of year-end all types of overdoses to the counts resulted from the model when we sequentially add 2013 fatal overdose count target to the calibration process. (C). Comparison of accepted vs rejected samples of no-treatment overdose multiplier from the calibration process corresponding to (B).

### Figure S4: Model External Validity for Age-gender Stratified Total OUD Counts

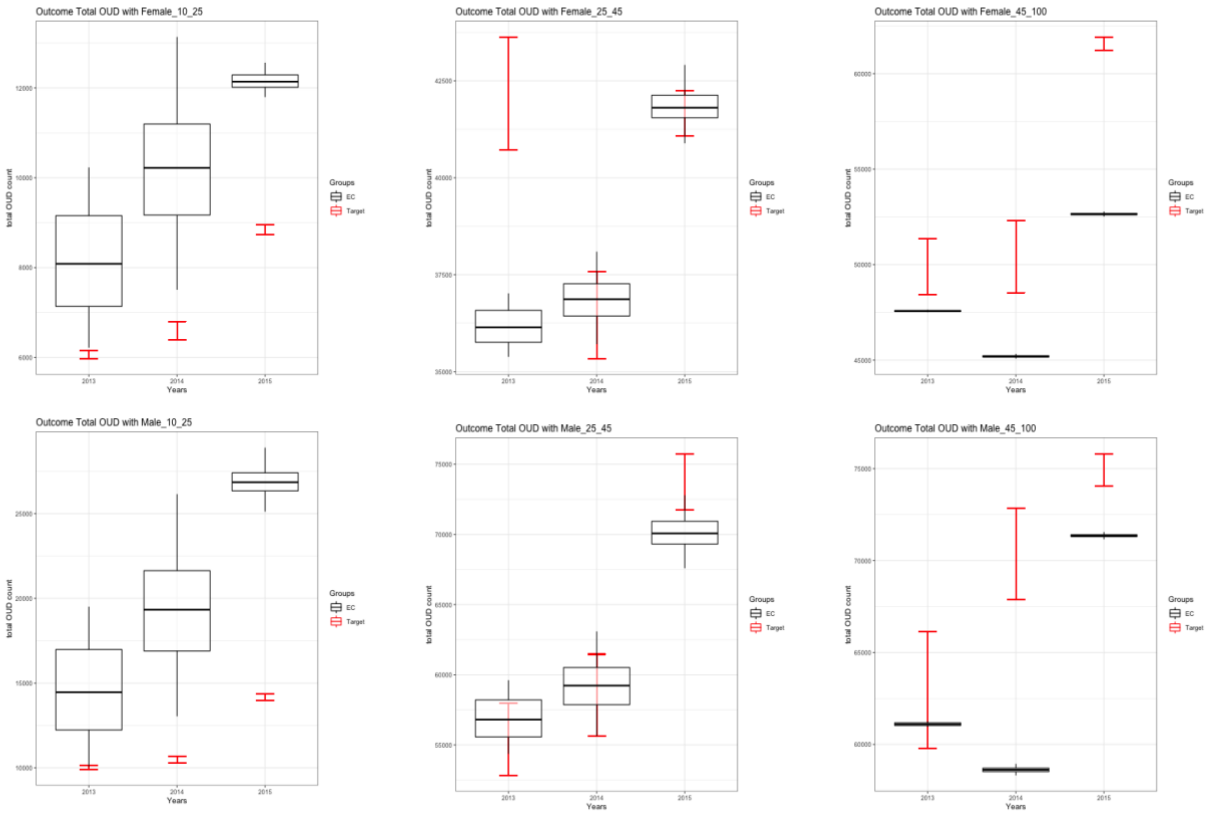


Figure S4 shows year 2015 age-gender stratified total OUD counts resulted from the calibrated model in comparison to targets estimated from the capture-recapture analysis.

#### Figure S5: Model External Validity for Age-gender Stratified Overdose Counts

Fatal Overdose Counts

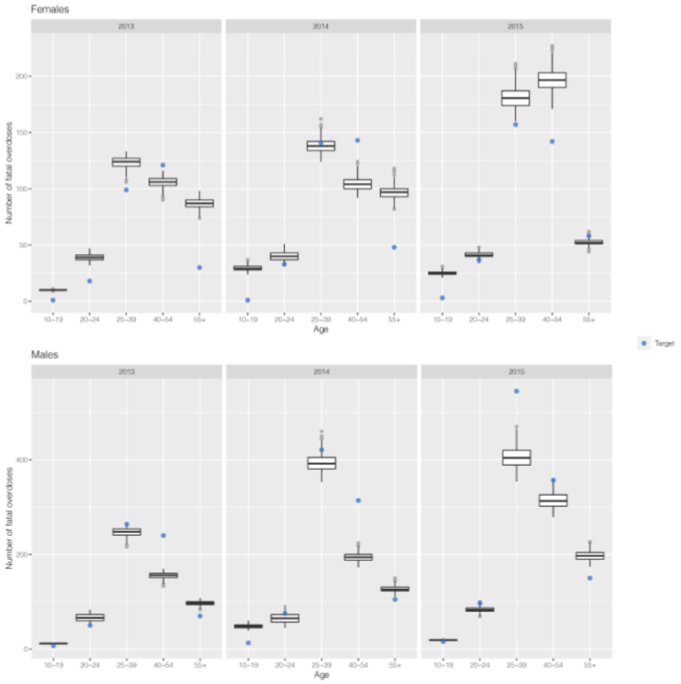


All Types of Overdoses

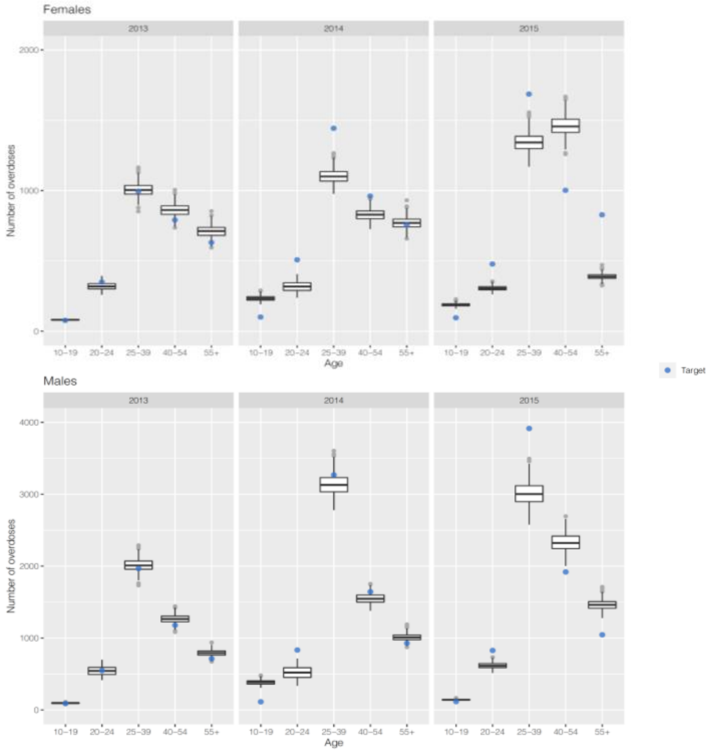


Figure S5 shows a comparison of age-gender stratified all types of overdose counts (both non-fatal and fatal) from the calibrated model to observed overdose count targets.

**Figure S6: Model Validity for Admissions to Treatment**

Buprenorphine

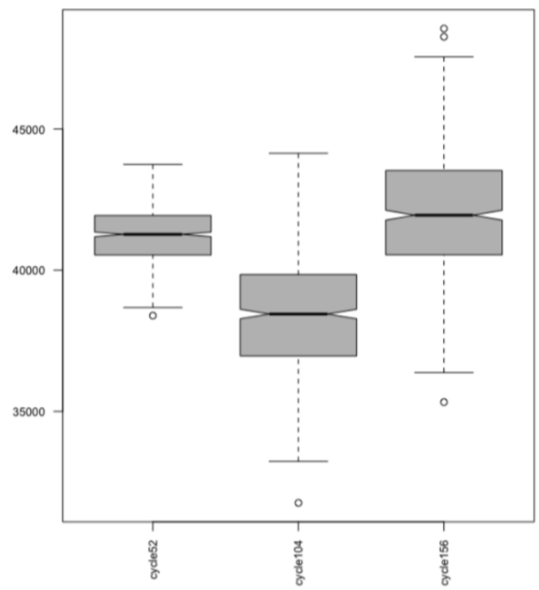


Naltrexone

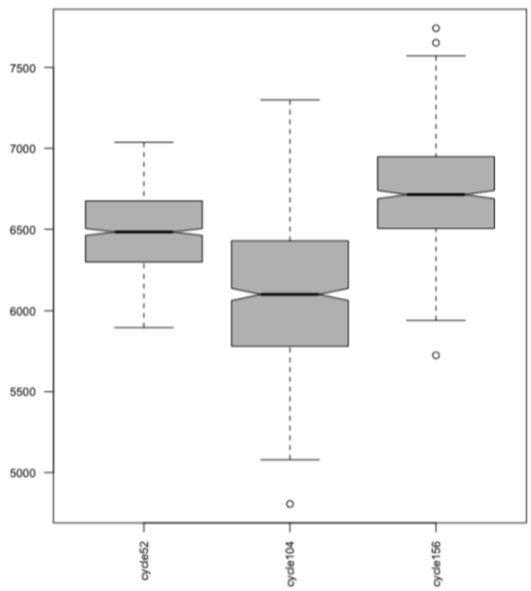


Methadone

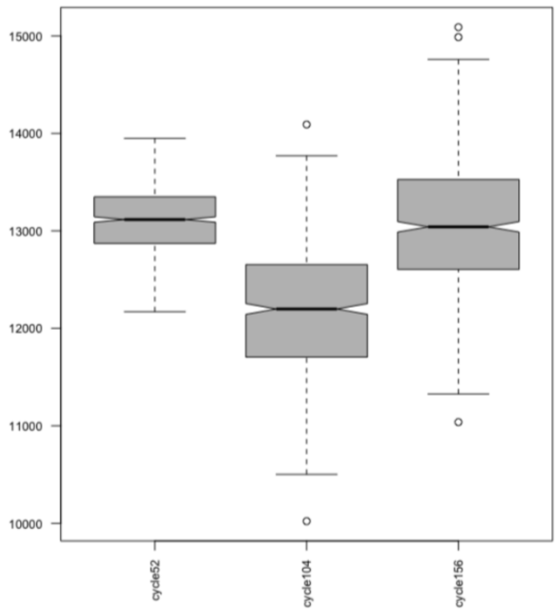


Figure S6 shows yearly MOUD admission counts resulted from the calibrated model.

**Table S1. Model Outcomes vs Calibration Targets**

| | Year | Observed (95% CI) | Median (2.5$^{th}$ percentile, 97.5$^{th}$ percentile) | Mean (95% CI) |
|---|---|---|---|---|
| Total OUD | 2013 | 226861 (222242, 231285)**, *** | 224168 (214904, 234799) | 224530 (224091, 224970) |
| | 2014 | 233184 (228680, 237514)**, *** | 229501 (217501, 241447) | 229397 (228914, 229880) |
| | 2015 | 275070 (272707, 277402) *** | 274949 (272781, 277294) | 274954 (274856, 275053) |
| Admissions to detox* | 2013 | 39635 | 39559 (35883, 43405) | 39614 (39452, 39775) |
| | 2014 | 41229 | 41178 (37289, 45086) | 41183 (41014, 41353) |
| | 2015 | 38329 | 38257 (34712, 41958) | 38256 (38094, 38417) |
| Fatal overdoses* | 2013 | 900 | 949 (862, 988) | 943 (940, 945) |
| | 2014 | 1294 | 1229 (1168, 1358) | 1238 (1234, 1242) |
| | 2015 | 1562 | 1508 (1415, 1666) | 1518 (1513, 1523) |

* These are observed counts from DPH with no known uncertainty around the values. Therefore, we use within 10% of the values as the uncertainty interval for the empirical calibration.
** We didn't use total OUD counts and their ranges of 2013 and 2014 directly as targets in empirical calibration. Values shown here are used only to create in Figure 1(A).
*** Estimated from capture-recapture analysis [1].

**Table S2. Alive OUD Percentages**

| | Year | Median (2.5th percentile, 97.5th percentile) |
|---|---|---|
| In no-treatment | 2013 | 80.99 (80.34, 81.46) |
| | 2014 | 82.48 (80.99, 83.37) |
| | 2015 | 84.31 (82.62, 85.44) |
| In all treatments | 2013 | 15.55 (15.14, 16.13) |
| | 2014 | 14.21 (13.31, 15.43) |
| | 2015 | 12.82 (11.84, 14.29) |
| In all post-treatments | 2013 | 3.45 (3.35, 3.60) |
| | 2014 | 3.32 (3.14, 3.58) |
| | 2015 | 2.91 (2.71, 3.10) |

**Table S3. Overall Active OUD Percentages Distributed among Health States**

| | Year | Median (2.5$^{th}$ percentile, 97.5$^{th}$ percentile) |
|---|---|---|
| In no-treatment | 2013 | 90.48 (90.15, 90.86) |
| | 2014 | 90.45 (90.02, 90.76) |
| | 2015 | 91.51 (91.06, 91.99) |
| In all treatments | 2013 | 5.88 (5.69, 6.07) |
| | 2014 | 5.81 (5.64, 5.94) |
| | 2015 | 5.23(5.09, 5.36) |
| In all post-treatments | 2013 | 3.64 (3.17, 4.09) |
| | 2014 | 3.73 (3.36, 4.31) |
| | 2015 | 3.24 (2.85, 3.68) |

**Table S4.** Active OUD Percentages

| | Year | Median (2.5$^{th}$ percentile, 97.5$^{th}$ percentile) |
|---|---|---|
| In no-treatment | 2013 | 80.51 (76.18, 86.03) |
| | 2014 | 72.39 (66.06, 82.89) |
| | 2015 | 72.10 (65.20, 83.19) |
| In all treatments | 2013 | 27.24 (27.13, 27.38) |
| | 2014 | 27.19 (26.94, 27.44) |
| | 2015 | 27.13 (26.98, 27.28) |
| In all post-treatments | 2013 | 74.21 (65.16, 89.72) |
| | 2014 | 74.03 (64.79, 89.87) |
| | 2015 | 73.95 (63.52, 89.86) |

**References**


[1] Barocas JA, White LF, Wang J, et al. Estimated Prevalence of Opioid Use Disorder in Massachusetts, 2011-2015: A Capture-Recapture Analysis. *Am J Public Health* 2018: e1-e7. DOI: 10.2105/AJPH.2018.304673.

[2] Morgan, J.R., et al., *Injectable naltrexone, oral naltrexone, and buprenorphine utilization and discontinuation among individuals treated for opioid use disorder in a United States commercially insured population.* J Subst Abuse Treat, 2018. **85**: p. 90-96.

[3] Sordo, L., et al., *Mortality risk during and after opioid substitution treatment: systematic review and meta-analysis of cohort studies.* BMJ, 2017. **357**: p. j1550.
